# Site-specific cancer incidence by race and immigration status in Canada 2006-2015: a population-based data linkage study

**DOI:** 10.1101/2022.11.09.22281329

**Authors:** Talía Malagón, Samantha Morais, Parker Tope, Mariam El-Zein, Eduardo L Franco

## Abstract

**Introduction:** The Canadian Cancer Registry does not collect demographic data beyond age and sex, making it hard to monitor health inequalities in cancer incidence in Canada, a country with public healthcare and many immigrants. Using data linkage, we compared site-specific cancer incidence rates by race.

**Methods:** We used data from the 2006 and 2011 Canadian Census Health and Environment Cohorts (CanCHECs), which are population-based probabilistically linked datasets of 5.9 million respondents of the 2006 Canadian long-form census and 6.5 million respondents of the 2011 National Household Survey. Respondents’ race was self-reported using Indigenous identity and visible minority group identity questions. Respondent data were linked with the Canadian Cancer Registry up to 2015. We calculated age-standardized incidence rate ratios (ASIRR), comparing group-specific rates to the overall population rate with bootstrapped 95% confidence intervals (95%CI). We used negative binomial regressions to adjust rates for socioeconomic variables and assess interactions with immigration status.

**Results:** The age-standardized cancer incidence rate was lower in almost all non-White racial groups than in White individuals, except for Indigenous peoples who had a similar overall age-standardized cancer incidence rate (ASIRR 0.99, 95%CI 0.97-1.01). Immigrants had substantially lower age-standardized overall cancer incidence rates than non-immigrants (ASIRR 0.83, 95%CI 0.82-0.84). Non-White racial groups generally had significantly lower or equivalent site-specific cancer incidence rates than the overall population, except for stomach, liver, and thyroid cancers and for multiple myeloma. Differences in incidence rates by race persisted even after adjusting for household income, education, and rural residence, with immigration status being an important modifier of cancer risk.

**Conclusions:** Differences in cancer incidence between racial groups are likely influenced by differences in lifestyles and early life exposures, as well as selection factors for immigration. This suggests a strong role of environment in determining cancer risk and further potential for cancer prevention.

## Introduction

The collection of race-based and Indigenous identity data is essential to measure and address health inequalities. Most evidence regarding racial inequalities in cancer health outcomes comes from the United States;^1,2^ however, findings from the United States may not be transposable to other health systems with different populations and contextual factors influencing health. The examination of race-based outcomes in other countries can help to identify commonalities and potential differences in drivers of cancer inequalities.

Canada is a country with a public healthcare system and universal health coverage. It is ethnically and racially diverse with a large proportion of immigrants. While evidence exists that cancer screening, incidence, and survival differ by race and ethnicity in Canada,^3-7^ recent national-level estimates of site-specific cancer incidence by race are lacking. The Canadian Cancer Registry does not collect demographic data beyond age and sex, making it difficult to monitor inequalities in cancer incidence and mortality across Canada. There is a legacy of colonialism and a historical reluctance to gather race-based data in Canada, which have led to a current paucity of data for monitoring racial health inequalities.^8,9^ There have been recent calls to provide more racially disaggregated health data.^1,8^ For historical and legal reasons, race data is often collected by Canadian institutions using the controversial^10^ construct of ‘visible minority group’, defined as persons, other than Indigenous peoples, who are non-Caucasian in race or non-white in colour. In 2022 the Canadian Institute of Health Information (CIHI) published updated guidance on the use of standards for race-based and Indigenous identity data health reporting, dropping the term ‘visible minority’.^11^ These standards distinguish between race and ethnicity, with race defined as a social construct used to categorize people based on perceived differences in physical appearance, and ethnicity defined as a multi-dimensional construct based on community belonging and a shared cultural group membership. This definition of race subsumes the construct of visible minority by including White and Indigenous populations. However, because much of the data collected in Canada still uses visible minority group definitions, in this paper we use the term race when referring to all racial groups and the term visible minority groups when referring specifically to racial groups who are not White and not Indigenous.

Due to the low incidence rate of some cancers, large data-linkage studies are required to monitor racial inequalities for many cancers in Canada. Our objective was to measure differences in site-specific cancer incidence across racial groups in a representative Canadian population to identify inequalities in cancer incidence. We also assessed whether socioeconomic status and immigration contribute to differences in cancer incidence by race.

## Methods

### Study sample and data sources

The study sample includes members of the 2006 and 2011 Canadian Census Health and Environment Cohorts (CanCHECs), a set of population-based probabilistically linked datasets based on the long-form census and the National Household Survey (NHS).^12^ The 2006 long-form census questionnaire was a mandatory questionnaire sent to approximately one in five Canadian households collecting information on relationships, languages, labour, income, education, housing, immigration, race, and ethnic origin.^13^ The long-form census was replaced with the NHS in 2011, a voluntary survey collecting the same information as the long-form census sent to approximately one in three Canadian households.^14^ Both surveys targeted the non-institutionalized usual residents of Canada on Census Day (mid-May 2006 or 2011), including both permanent and non-permanent residents. Survey respondents were probabilistically linked using personal identifiers to the Derived Record Depository (DRD), a national dynamic relational database of individuals residing in Canada.^15^ The 2006 and 2011 CanCHEC cohorts include the 90.8% of the 2006 long-form census respondents and 96.7% of 2011 NHS respondents who could be linked to the DRD.^12^

Cohort records were linked via the DRD up to December 31^st^ 2015 to the Canadian Vital Statistics Death Database (CVSD)^16^, the Canadian Cancer Registry (CCR)^17^, and the annual postal codes file^18^. The CVSD collects dates and cause of death information annually from all provincial and territorial vital statistics registries on all deaths in Canada. The CCR is a population-based registry that includes information about each new primary cancer diagnosed in Canada. The International Agency for Research on Cancer (IARC) rules were used for determining multiple primary cancers in the CCR.^19^ Cancers were grouped by site using Surveillance, Epidemiology, and End Results (SEER) site groupings based on ICD-O-3 codes (Supplementary Table 1). For head and neck cancers, we used the definition used by the 2021 Canadian Cancer Statistics.^20^

The annual postal code file collects the mailing addresses provided on income tax returns by tax filers. Postal codes were used to estimate changes in a person’s place of residence for each year of follow-up after the census year (2006 or 2011). For individuals in the CCR or CVSD, we used the residential postal code available in these registries instead of the annual postal code file for the years in which they were diagnosed with cancer or died. The geographical information related to each postal code was extracted using the Postal Code Conversion File Plus program version 7D.^21^ Postal codes were not available for all cohort members or for all follow-up years due to missing tax records or census respondents who did not give permission to link tax returns. For years with missing postal code data, we imputed the last known place of residence from the CCR, annual postal code file, long-form census/NHS (in order of priority) forward to subsequent years.

### Racial group & immigrant status

Racial identity and immigrant status were self-reported based on the Aboriginal group, visible minority group, immigrant status, and citizenship questions in the long-form census (2006) and the NHS (2011).^22,23^ The racial categories in these questions are based on the visible minority group definitions used for federal employment equity programs in Canada. The visible minority group question was directed to respondents who did not identify as Indigenous, and included 11 response categories (White, South Asian, Chinese, Black, Filipino, Latin American, Arab, Southeast Asian, West Asian, Korean, and Japanese) and a write-in space. Respondents who reported belonging only to a single group were included in that corresponding racial group category. Respondents who selected multiple groups were classified using rules used by employment equity definitions.^22^ Respondents who reported being both ‘White’ and ‘Chinese’, ‘South Asian’, ‘Black’, ‘Filipino’, ‘Southeast Asian’, ‘Japanese’, or ‘Korean’ were included in their corresponding visible minority group category. Respondents who reported being both ‘White’ and ‘Latin American’, ‘Arab’, or ‘West Asian’ were included in the ‘White’ category, as these are not considered to be visible minority groups by employment equity definitions and are grouped with ‘White’ in survey data products. Respondents who reported belonging to multiple non-White visible minority groups were categorized as ‘Multiple visible minority groups’. Those who provided a write-in response not classifiable into any other group were categorized as ‘Other visible minority groups’. Native-born Canadians (non-immigrants) are those who indicated they were born in Canada; immigrants are those born outside of Canada who have been granted the right to live in Canada permanently by immigration authorities; and non-permanent residents are those who are neither born in Canada nor immigrants (ex. person with a temporary work/study permit).

### Statistical analyses

The 2006 and 2011 CanCHECs were pooled together to increase the sample size and statistical power for the analysis. Respondents contributed person-time at risk and events from the year of their survey response (2006 or 2011) up to their year of death or December 31^st^ 2015. Because the province of Québec has not reported new cancer cases to the CCR since 2010, we excluded person-time from Québec residents after 2010. To calculate person-time and events by age, we used each person’s age at the midpoint of each calendar year (July 1^st^) of follow-up.

We calculated age-standardized cancer incidence rates and age-standardized incidence rate ratios (ASIRR) by population group using the 2011 Canadian population age distribution as standard.^24^ Age-specific incidence rates were weighted by CanCHECs survey weights in order to be representative of the Canadian population living in private dwellings. The CanCHECs survey weights are calibrated to account for survey sampling probability, non-response rates, and DRD linkage probabilities.^12^ The 95% confidence intervals (CI) for age-standardized cancer incidence rates and ASIRR were calculated through bootstrapping, using the 2.5^th^ and 97.5^th^ percentile intervals of 500 bootstrap replicate weights developed for the CanCHECs. To validate sample representativeness, we compared cancer incidence rates in the CanCHECs with national cancer incidence rates reported during the same period; the results were highly similar (Supplementary Tables 2-3). In secondary analyses, we also age-standardized using the 1960 World Population for comparisons with cancer incidence rates in the world region of origin of visible minority groups using GLOBOCAN 2020 data.^25,26^

To assess whether incidence rate ratios between racial groups could be attributed to a healthy immigrant effect or differences in socioeconomic status, we used negative binomial regression models. Regression models used the logarithm of group person-years at risk as an offset. These regression models included an interaction term between race and immigration status to calculate different rates for native-born Canadians compared with foreign-born immigrants and non-permanent residents. Incidence rates were adjusted for 5-year age group, sex, calendar year, province of residence, rural/urban place of residence, education level, and quintiles of household after-tax income adjusted for household size. Due to the small numbers of events in some racial groups when assessing interactions with immigration, some populations were grouped together in regression analyses to improve model stability while maintaining meaningful distinctions for race-based reporting: Chinese, Filipino, Southeast Asian, Korean, and Japanese were grouped as East/Southeast Asian; West Asian and Arab were grouped as Middle Eastern; and those in multiple visible minority groups were combined with the other visible minority groups category.

### Ethics and confidentiality

We obtained ethical approval from the McGill University Institutional Review Board for this analysis of secondary data. To protect respondent confidentiality, analysis outputs were vetted using rules developed by Statistics Canada, which include rounding of all counts to the nearest 5 and not disclosing descriptive statistics for groups with less than 5 events. In analyses where the number of cancer cases in a racial group did not meet the disclosure threshold, we either combined this group with the ‘Other visible minority group’ or suppressed outputs as indicated.

## Results

There were 5.9 million respondents in the 2006 CanCHEC and 6.5 million respondents in the 2011 CanCHEC (Table 1). The most common racial groups in both cohorts were White (75-78%), Indigenous peoples (7-8%), South Asian (4-5%), and Chinese (3-4%) populations. Immigrants constituted 18-20% of respondents and non-permanent residents 1% of respondents. The proportion of immigrants and non-permanent residents was substantially higher among visible minority groups than among White individuals and Indigenous peoples. The median time since immigration for immigrants was 18 years (interquartile range 8-36).

**Table 1.**
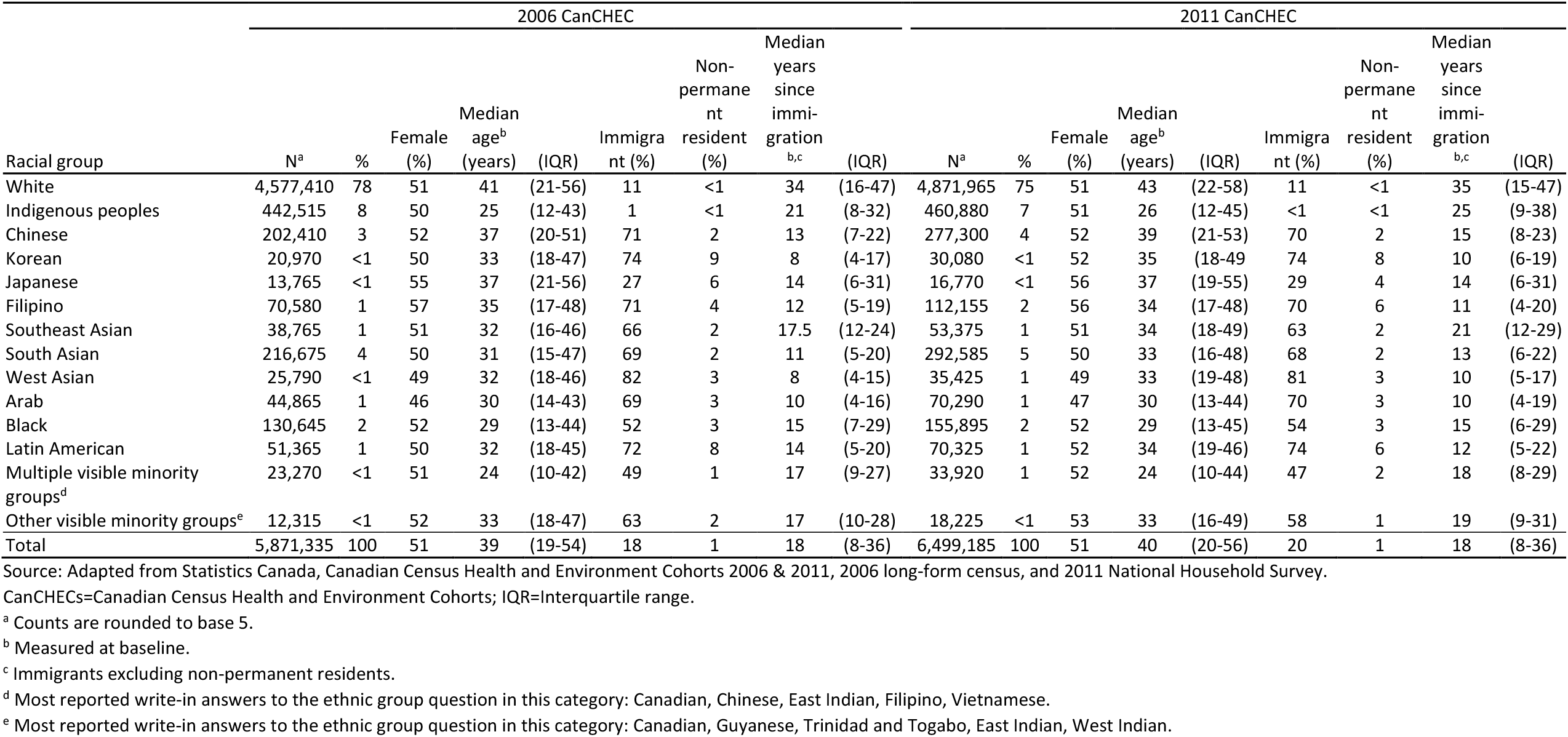
Baseline characteristics of 2006 and 2011 CanCHECs by race.

| Racial group | 2006 CanCHEC |  |  |  |  |  |  |  |  | 2011 CanCHEC |  |  |  |  |  |  |  |  |
| --- | --- | --- | --- | --- | --- | --- | --- | --- | --- | --- | --- | --- | --- | --- | --- | --- | --- | --- |
|  | N<sup>a</sup> | % | Female (%) | Median age<sup>b</sup> (years) | (IQR) | Immigrant (%) | Non-permanent resident (%) | Median years since immigration<sup>b,c</sup> | (IQR) | N<sup>a</sup> | % | Female (%) | Median age<sup>b</sup> (years) | (IQR) | Immigrant (%) | Non-permanent resident (%) | Median years since immigration<sup>b,c</sup> | (IQR) |
| White | 4,577,410 | 78 | 51 | 41 | (21-56) | 11 | <1 | 34 | (16-47) | 4,871,965 | 75 | 51 | 43 | (22-58) | 11 | <1 | 35 | (15-47) |
| Indigenous peoples | 442,515 | 8 | 50 | 25 | (12-43) | 1 | <1 | 21 | (8-32) | 460,880 | 7 | 51 | 26 | (12-45) | <1 | <1 | 25 | (9-38) |
| Chinese | 202,410 | 3 | 52 | 37 | (20-51) | 71 | 2 | 13 | (7-22) | 277,300 | 4 | 52 | 39 | (21-53) | 70 | 2 | 15 | (8-23) |
| Korean | 20,970 | <1 | 50 | 33 | (18-47) | 74 | 9 | 8 | (4-17) | 30,080 | <1 | 52 | 35 | (18-49) | 74 | 8 | 10 | (6-19) |
| Japanese | 13,765 | <1 | 55 | 37 | (21-56) | 27 | 6 | 14 | (6-31) | 16,770 | <1 | 56 | 37 | (19-55) | 29 | 4 | 14 | (6-31) |
| Filipino | 70,580 | 1 | 57 | 35 | (17-48) | 71 | 4 | 12 | (5-19) | 112,155 | 2 | 56 | 34 | (17-48) | 70 | 6 | 11 | (4-20) |
| Southeast Asian | 38,765 | 1 | 51 | 32 | (16-46) | 66 | 2 | 17.5 | (12-24) | 53,375 | 1 | 51 | 34 | (18-49) | 63 | 2 | 21 | (12-29) |
| South Asian | 216,675 | 4 | 50 | 31 | (15-47) | 69 | 2 | 11 | (5-20) | 292,585 | 5 | 50 | 33 | (16-48) | 68 | 2 | 13 | (6-22) |
| West Asian | 25,790 | <1 | 49 | 32 | (18-46) | 82 | 3 | 8 | (4-15) | 35,425 | 1 | 49 | 33 | (19-48) | 81 | 3 | 10 | (5-17) |
| Arab | 44,865 | 1 | 46 | 30 | (14-43) | 69 | 3 | 10 | (4-16) | 70,290 | 1 | 47 | 30 | (13-44) | 70 | 3 | 10 | (4-19) |
| Black | 130,645 | 2 | 52 | 29 | (13-44) | 52 | 3 | 15 | (7-29) | 155,895 | 2 | 52 | 29 | (13-45) | 54 | 3 | 15 | (6-29) |
| Latin American | 51,365 | 1 | 50 | 32 | (18-45) | 72 | 8 | 14 | (5-20) | 70,325 | 1 | 52 | 34 | (19-46) | 74 | 6 | 12 | (5-22) |
| Multiple visible minority groups<sup>d</sup> | 23,270 | <1 | 51 | 24 | (10-42) | 49 | 1 | 17 | (9-27) | 33,920 | 1 | 52 | 24 | (10-44) | 47 | 2 | 18 | (8-29) |
| Other visible minority groups<sup>e</sup> | 12,315 | <1 | 52 | 33 | (18-47) | 63 | 2 | 17 | (10-28) | 18,225 | <1 | 53 | 33 | (16-49) | 58 | 1 | 19 | (9-31) |
| Total | 5,871,335 | 100 | 51 | 39 | (19-54) | 18 | 1 | 18 | (8-36) | 6,499,185 | 100 | 51 | 40 | (20-56) | 20 | 1 | 18 | (8-36) |

Source: Adapted from Statistics Canada, Canadian Census Health and Environment Cohorts 2006 & 2011, 2006 long-form census, and 2011 National Household Survey.
CanCHECs=Canadian Census Health and Environment Cohorts; IQR=Interquartile range.
<sup>a</sup> Counts are rounded to base 5.
<sup>b</sup> Measured at baseline.
<sup>c</sup> Immigrants excluding non-permanent residents.
<sup>d</sup> Most reported write-in answers to the ethnic group question in this category: Canadian, Chinese, East Indian, Filipino, Vietnamese.
<sup>e</sup> Most reported write-in answers to the ethnic group question in this category: Canadian, Guyanese, Trinidad and Tobago, East Indian, West Indian.

Age-standardized cancer incidence rates by race, sex, and immigration status for all cancer sites combined are presented in Table 2 and Figure 1. Age-standardized cancer incidence rates were lower in nearly all non-White racial groups than in White individuals, except for Indigenous peoples who had a similar overall age-standardized cancer incidence rate (ASIRR 0.99, 95%CI 0.97-1.01). Overall, women had a lower age-standardized overall cancer incidence rate than men (ASIRR 0.83, 95%CI 0.82-0.83), but the female to male cancer ASIRR displayed substantial variation by racial group. Immigrants had substantially lower age-standardized overall cancer incidence rates than native-born Canadians (ASIRR 0.83, 95%CI 0.82-0.84). The ASIRR between immigrant and native-born Canadians also displayed substantial variation by racial group.

**Table 2.** Age-standardized cancer incidence rate for all sites combined per 100,000 person-years and incidence rate ratios by race, age-standardized to the Canadian 2011 census population.

| Racial group | Events<sup>a</sup> | Age-standardized cancer incidence rates (/100,000 person-years) |  |  |  |  |  | Age-standardized incidence rate ratios |  |  |  |  |  |
| --- | --- | --- | --- | --- | --- | --- | --- | --- | --- | --- | --- | --- | --- |
|  |  | Overall | Females | Males | Native-born | Immigrants | Non-permanent residents | Racial group vs. White<br>(Reference) | (95% CI) | Females vs. males | (95% CI) | Immigrants vs. Native-born | (95% CI) |
| Overall | 411,220 | 514 | 471 | 570 | 542 | 450 | 392 | - | - | 0.83 | (0.82-0.83) | 0.83 | (0.82-0.84) |
| White | 355,555 | 536 | 488 | 599 | 544 | 512 | 447 | (Reference) | - | 0.81 | (0.81-0.82) | 0.94 | (0.93-0.95) |
| Indigenous peoples | 17,950 | 529 | 507 | 559 | 530 | 472 | 730 | 0.99 | (0.97-1.01) | 0.91 | (0.86-0.95) | 0.89 | (0.70-1.11) |
| Chinese | 11,455 | 360 | 361 | 360 | 392 | 358 | 308 | 0.67 | (0.66-0.68) | 1.00 | (0.96-1.05) | 0.91 | (0.83-1.02) |
| Korean | 950 | 342 | 319 | 367 | 516 | 341 | 337 | 0.64 | (0.58-0.69) | 0.87 | (0.74-1.01) | 0.66 | (0.42-1.47) |
| Japanese | 975 | 413 | 426 | 394 | 408 | 404 | 218 | 0.77 | (0.72-0.83) | 1.08 | (0.93-1.25) | 0.99 | (0.85-1.17) |
| Filipino | 3,915 | 407 | 402 | 425 | 439 | 407 | 334 | 0.76 | (0.73-0.79) | 0.94 | (0.88-1.04) | 0.93 | (0.63-1.34) |
| Southeast Asian | 1,290 | 326 | 310 | 352 | 451 | 319 | 300 | 0.61 | (0.57-0.65) | 0.88 | (0.77-1.03) | 0.71 | (0.51-1.08) |
| South Asian | 8,195 | 297 | 308 | 289 | 394 | 294 | 200 | 0.55 | (0.54-0.57) | 1.06 | (1.01-1.12) | 0.74 | (0.59-0.95) |
| West Asian | 1,090 | 379 | 343 | 415 | 644 | 374 | 250 | 0.71 | (0.65-0.75) | 0.83 | (0.73-0.96) | 0.58 | (0.38-1.00) |
| Arab | 1,480 | 445 | 442 | 450 | 626 | 432 | 625 | 0.83 | (0.77-0.89) | 0.98 | (0.86-1.12) | 0.69 | (0.53-0.91) |
| Black | 5,225 | 451 | 373 | 552 | 588 | 434 | 420 | 0.84 | (0.81-0.87) | 0.68 | (0.63-0.73) | 0.74 | (0.67-0.82) |
| Latin American | 1,615 | 353 | 331 | 390 | 436 | 353 | 309 | 0.66 | (0.61-0.70) | 0.85 | (0.74-0.97) | 0.81 | (0.54-1.37) |
| Multiple visible minority groups | 905 | 416 | 422 | 416 | 574 | 403 | 477 | 0.78 | (0.72-0.84) | 1.01 | (0.86-1.20) | 0.70 | (0.49-1.04) |
| Other visible minority groups | 625 | 397 | 371 | 447 | 404 | 398 | 194 | 0.74 | (0.67-0.82) | 0.83 | (0.66-0.99) | 0.99 | (0.61-2.18) |

Source: Adapted from Statistics Canada, Canadian Census Health and Environment Cohorts 2006 & 2011, 2006 long-form census, 2011 National Household Survey, Canadian Vital Statistics Death Database 2006-2015, and Canadian Cancer Registry 2006-2015.
95% CI=confidence intervals based on 2.5<sup>th</sup> and 97.5<sup>th</sup> bootstrap percentiles
<sup>a</sup> Counts are rounded to base 5.

**Figure 1.**
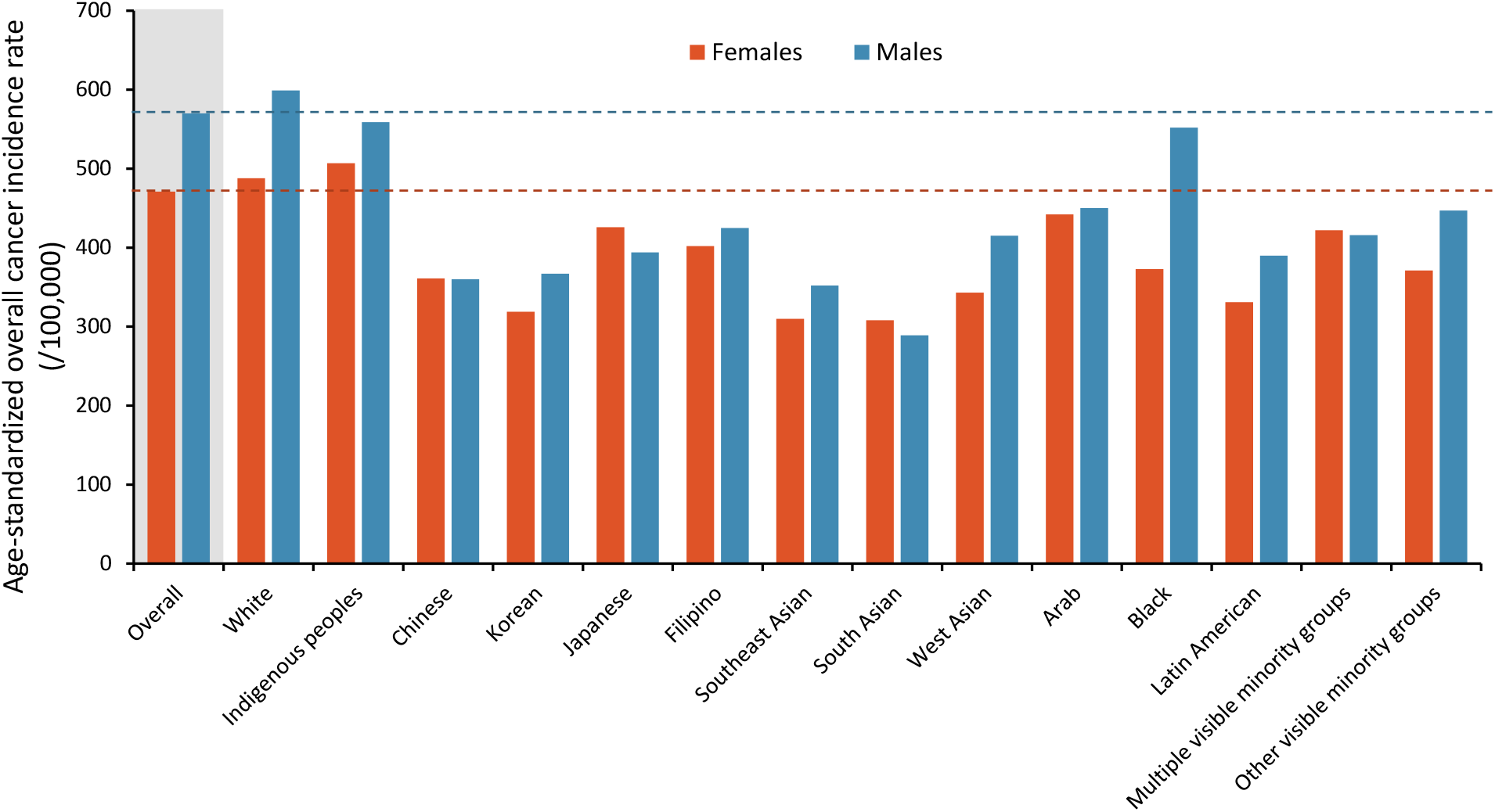
Age-standardized incidence rate of cancer per 100,000 person-years by sex and race, all cancer sites combined. Age-standardized to the 2011 Canadian census population. Source: Adapted from Statistics Canada, Canadian Census Health and Environment Cohorts 2006 & 2011, 2006 long-form census, 2011 National Household Survey, Canadian Vital Statistics Death Database 2006-2015, and the Canadian Cancer Registry 2006-2015.

Site-specific age-standardized incidence rates and ASIRRs are presented in Table 3 and Table 4, respectively. White individuals had higher age-standardized incidence rates than the overall population across most sites, with the notable exceptions of stomach, liver, and thyroid cancers, and multiple myelomas. With these cancers excepted, visible minority groups generally had lower or equivalent age-standardized cancer incidence rates compared to the overall population. Indigenous peoples had higher incidence rates for certain cancer sites (head and neck, stomach, colorectal, liver, pancreas, lung and bronchus, cervical, kidney and renal pelvis) but lower incidence rates for other cancer sites (melanoma, uterus, prostate, testis, bladder, brain and central nervous system, thyroid, Hodgkin Lymphoma, and leukemia) compared to the overall population.

**Table 3.** Age-standardized cancer incidence rates per 100,000 person-years by cancer site and race, age-standardized to the Canadian 2011 census population.

| Cancer site | Total number of events<sup>a</sup> | Age-standardized cancer incidence rates /100,000 person-years (95% CI) |  |  |  |  |  |  |  |  |  |  |  |  | Multiple visible minority groups | Other visible minority groups<sup>b</sup> |
| --- | --- | --- | --- | --- | --- | --- | --- | --- | --- | --- | --- | --- | --- | --- | --- | --- |
|  |  | Overall | White | Indigenous peoples | Chinese | Korean | Japanese | Filipino | Southeast Asian | South Asian | West Asian | Arab | Black | Latin American |  |  |
| All cancers | 411,220 | 514<br>(512-516) | 536<br>(534-538) | 529<br>(518-544) | 360<br>(352-367) | 342<br>(313-371) | 413<br>(385-445) | 407<br>(392-422) | 326<br>(303-350) | 297<br>(289-304) | 379<br>(348-404) | 445<br>(415-475) | 451<br>(434-464) | 353<br>(329-374) | 416<br>(384-451) | 397<br>(360-441) |
| Head and neck | 12,315 | 15.4<br>(15.0-15.7) | 16.1<br>(15.7-16.5) | 18.6<br>(16.5-20.9) | 12.2<br>(10.9-13.5) | 2.6<br>(0.8-4.6) | 8.7<br>(4.7-13.1) | 10.6<br>(8.1-12.8) | 7.4<br>(4.0-10.3) | 10.9<br>(9.5-12.5) | 9.6<br>(6.2-13.4) | 6.6<br>(3.9-9.8) | 6.6<br>(4.7-8.6) | 8.0<br>(5.1-12.1) | 13.2<br>(6.7-19.0) | 11.9<br>(6.0-18.6) |
| Esophagus | 4,125 | 5.2<br>(5.0-5.4) | 5.7<br>(5.4-5.9) | 6.1<br>(4.8-7.4) | 1.8<br>(1.2-2.2) | <sup>c</sup> | 5.8<br>(2.0-8.1) | 0.9<br>(0.3-1.6) | <sup>c</sup> | 3.1<br>(2.3-4.1) | 2.4<br>(0.6-4.4) | <sup>c</sup> | 2.8<br>(1.5-4.3) | 0.9<br>(0.2-1.6) | <sup>c</sup> | 1.6<br>(0.7-2.5) |
| Stomach | 7,435 | 9.3<br>(9.0-9.5) | 9.0<br>(8.7-9.3) | 11.1<br>(9.4-12.9) | 10.7<br>(9.4-11.9) | 30.6<br>(23.8-37.5) | 26.1<br>(18.0-35.0) | 6.9<br>(4.8-9.2) | 11<br>(7.2-15.0) | 5.9<br>(4.7-7.2) | 9.9<br>(5.3-14.8) | 14.5<br>(9.4-19.8) | 12<br>(9.3-14.3) | 17.4<br>(12.3-22.7) | 9.9<br>(5.3-14.8) | 10.9<br>(5.8-16.3) |
| Colorectal | 50,595 | 62.4<br>(61.8-63.0) | 64.9<br>(64.1-65.5) | 79.6<br>(75.0-85.0) | 50<br>(47.0-53.0) | 41.2<br>(33.0-51.1) | 68.2<br>(58.3-81.0) | 47.4<br>(42.2-53.9) | 43.1<br>(35.0-51.4) | 26.5<br>(24.2-28.5) | 39.7<br>(29.5-48.9) | 47.4<br>(37.9-57.3) | 44.3<br>(39.7-48.9) | 36.5<br>(29.8-43.6) | 51.9<br>(40.9-64.2) | 44.5<br>(33.5-56.9) |
| Anal | 1,440 | 1.8<br>(1.7-2.0) | 2.0<br>(1.9-2.2) | 1.9<br>(1.1-2.7) | 0.3<br>(0.1-0.5) | <sup>c</sup> | <sup>c</sup> | 1.2<br>(0.4-2.0) | <sup>c</sup> | 0.7<br>(0.3-1.2) | <sup>c</sup> | <sup>c</sup> | 1.6<br>(0.9-2.5) | <sup>c</sup> | <sup>c</sup> | 0.7<br>(0.4-1.0) |
| Liver | 4,095 | 5.1<br>(4.9-5.3) | 4.4<br>(4.2-4.6) | 11.0<br>(8.9-13.3) | 12.9<br>(11.5-14.4) | 14.7<br>(10.0-20.3) | 5.1<br>(1.9-8.5) | 7.4<br>(5.5-9.4) | 20.2<br>(15.0-25.5) | 4.7<br>(3.7-5.7) | 2.6<br>(1.0-4.3) | 4.5<br>(2.2-7.1) | 5.1<br>(3.8-6.6) | 7.0<br>(4.0-10.2) | 13.4<br>(7.6-19.7) | 4.3<br>(1.7-7.7) |
| Pancreas | 9,885 | 12.4<br>(12.1-12.7) | 12.9<br>(12.6-13.2) | 14.3<br>(12.5-16.4) | 8.4<br>(7.2-9.5) | 11.2<br>(6.7-16.5) | 10.8<br>(6.9-14.8) | 9.1<br>(6.8-11.5) | 7.4<br>(4.2-10.6) | 6.9<br>(5.8-8.1) | 5.7<br>(2.1-9.7) | 8.2<br>(5.1-12.4) | 13.3<br>(10.4-16.2) | 10.6<br>(6.4-13.7) | 11.2<br>(6.1-16.6) | 5.5<br>(1.2-9.1) |
| Lung & bronchus | 52,990 | 66.0<br>(65.3-66.7) | 70.5<br>(69.7-71.3) | 89.7<br>(84.0-95.3) | 44.3<br>(41.2-46.7) | 38.4<br>(29.7-48.1) | 35.2<br>(27.2-43.2) | 39.3<br>(35.0-44.0) | 36.4<br>(27.6-43.8) | 14.1<br>(12.3-15.9) | 29.7<br>(19.9-38.1) | 32.0<br>(24.1-40.2) | 27.3<br>(23.6-31.1) | 26.4<br>(19.2-32.5) | 35.1<br>(25.7-43.9) | 27.1<br>(15.7-35.6) |
| Melanoma | 15,400 | 19.3<br>(19.0-19.7) | 22.9<br>(22.5-23.3) | 6.6<br>(5.4-8.0) | 1.2<br>(0.8-1.7) | <sup>c</sup> | <sup>c</sup> | 0.9<br>(0.4-1.6) | 2.0<br>(0.7-3.5) | 1.1<br>(0.8-1.6) | 1.6<br>(0.4-2.9) | 4.6<br>(2.5-7.3) | 3.3<br>(1.3-5.3) | 5.7<br>(2.9-7.7) | 2.1<br>(0.5-4.4) | 1.2<br>(0.3-2.1) |
| Breast (female) | 53,140 | 127.1<br>(125.8-128.4) | 130.2<br>(128.6-131.6) | 126.6<br>(118.3-134.8) | 107.1<br>(101.4-112.3) | 83.0<br>(68.4-100.8) | 142.9<br>(123.9-165.4) | 132<br>(121.5-140.2) | 75.3<br>(62.3-91.1) | 97.5<br>(92.3-102.8) | 121.5<br>(99.1-138.3) | 134.1<br>(113.7-154.1) | 102.0<br>(92.1-111.6) | 87.5<br>(72.8-103.1) | 142.7<br>(120.0-166.5) | 109.5<br>(85.9-135.4) |
| Cervix (female) | 3,045 | 7.4<br>(7.1-7.7) | 7.7<br>(7.4-8.1) | 12.0<br>(10.3-13.6) | 4.8<br>(3.7-5.9) | 6.3<br>(3.0-10.2) | 6.9<br>(2.8-12.0) | 8.2<br>(5.3-10.4) | 11.5<br>(7.1-16.3) | 4.9<br>(3.6-6.4) | <sup>c</sup> | 11.2<br>(1.1-18.6) | 6.9<br>(4.7-9.4) | 7.8<br>(4.6-11.9) | 9.8<br>(4.1-15.9) | 6.1<br>(2.1-9.8) |
| Uterus (female) | 13,080 | 31<br>(30.4-31.7) | 31.8<br>(31.1-32.6) | 23.1<br>(20.2-26.1) | 23.2<br>(20.7-25.7) | 12.5<br>(6.3-18.8) | 31.9<br>(22.0-42.7) | 35.5<br>(30.3-41.2) | 18.7<br>(12.5-25.6) | 29.3<br>(26.2-32.5) | 27.6<br>(17.7-38.8) | 23.9<br>(14.1-34.6) | 27.4<br>(22.5-33.0) | 27.1<br>(20.2-35.2) | 37.3<br>(20.5-56.8) | 20.8<br>(12.7-29.2) |
| Ovary (female) | 6,000 | 14.2<br>(13.7-14.6) | 14.4<br>(13.9-14.8) | 13.5<br>(11.1-16.9) | 11.2<br>(9.1-13.1) | 5.1<br>(2.2-8.4) | 22.7<br>(13.1-32.5) | 10.1<br>(7.7-12.3) | 15.9<br>(10.3-22.0) | 15.2<br>(12.7-17.7) | 14.2<br>(7.7-22.0) | 13.9<br>(6.6-21.2) | 11.6<br>(8.5-14.5) | 14.6<br>(7.8-22.2) | 10.9<br>(5.6-16.9) | 18.5<br>(5.4-33.1) |
| Prostate (male) | 52,840 | 138.5<br>(137.3-140.0) | 143.6<br>(142.2-145.1) | 114.4<br>(106.9-123.5) | 82<br>(77.3-87.5) | 76.2<br>(58.3-95.2) | 113<br>(92.5-135.8) | 123.2<br>(108.0-136.7) | 71.1<br>(55.6-87.9) | 79.4<br>(73.4-84.3) | 118.4<br>(96.9-138.8) | 97.1<br>(79.6-114.4) | 264.6<br>(247.2-279.5) | 111.2<br>(95.5-128.8) | 130.2<br>(103.9-161.3) | 129.8<br>(101.7-163.7) |
| Testis (male) | 2,120 | 5.9<br>(5.6-6.2) | 6.9<br>(6.5-7.2) | 4.6<br>(3.7-5.6) | 2.1<br>(1.5-2.9) | <sup>c</sup> | <sup>c</sup> | 1.2<br>(0.2-2.5) | <sup>c</sup> | 3.3<br>(2.5-4.2) | 5.8<br>(1.8-9.6) | 4.2<br>(1.9-6.7) | 1.9<br>(0.7-2.9) | 4.4<br>(2.6-6.5) | 2.3<br>(0.5-4.6) | 1.3<br>(0.6-2.2) |
| Bladder | 19,225 | 24.0<br>(23.6-24.4) | 26.1<br>(25.7-26.6) | 16.0<br>(13.8-18.2) | 9.5<br>(8.3-10.8) | 20.0<br>(12.7-27.1) | 10.5<br>(6.7-14.4) | 8.5<br>(6.0-10.8) | 5.3<br>(2.4-8.1) | 9.5<br>(8.1-10.9) | 18.6<br>(12.7-24.6) | 30.3<br>(22.2-37.5) | 10.2<br>(8.0-12.5) | 8.9<br>(5.2-12.7) | 9.0<br>(4.8-13.9) | 12.2<br>(6.7-18.5) |
| Kidney & renal<br>pelvis | 12,490 | 15.4<br>(15.0-15.7) | 15.9<br>(15.6-16.3) | 24.7<br>(22.3-27.2) | 8.5<br>(7.4-9.5) | 6.6<br>(3.5-9.4) | 10.9<br>(7.1-16.0) | 8.6<br>(6.5-10.6) | 11.1<br>(6.6-15.5) | 8.7<br>(7.4-10.1) | 11.4<br>(7.9-15.3) | 16.9<br>(11.1-23.5) | 10.7<br>(8.5-12.9) | 19.6<br>(15.0-24.6) | 7.9<br>(4.7-12.0) | 13.7<br>(6.8-21.4) |
| Brain & CNS | 5,725 | 7.5<br>(7.3-7.7) | 8.1<br>(7.9-8.4) | 5.0<br>(3.8-6.3) | 3.7<br>(2.9-4.4) | 3.3<br>(1.3-5.3) | <sup>c</sup> | 3.1<br>(2.0-4.3) | 5<br>(2.8-7.2) | 6.5<br>(5.3-7.6) | 5.6<br>(2.8-8.7) | 6.3<br>(3.0-9.7) | 4.5<br>(3.4-5.8) | 2.7<br>(1.6-4.1) | 5.2<br>(1.9-8.8) | 2.5<br>(1.0-4.1) |
| Thyroid | 12,370 | 16.1<br>(15.8-16.5) | 14.8<br>(14.5-15.2) | 10<br>(8.2-11.9) | 21<br>(19.5-22.7) | 30.2<br>(23.4-37.4) | 10.0<br>(5.7-14.4) | 37.3<br>(32.9-41.7) | 21.0<br>(16.5-25.6) | 19.9<br>(18.3-21.8) | 29.7<br>(23.1-35.8) | 30.9<br>(24.4-37.4) | 20.9<br>(18.0-23.5) | 19.5<br>(16.0-23.0) | 25.5<br>(19.0-32.4) | 34.8<br>(21.6-48.0) |
| Hodgkin<br>Lymphoma | 2,065 | 2.8<br>(2.7-3.0) | 3.1<br>(2.9-3.2) | 1.7<br>(1.1-2.3) | 1.4<br>(1.0-1.9) | <sup>c</sup> | <sup>c</sup> | 1<br>(0.4-1.7) | 1.7<br>(0.3-3.4) | 2.7<br>(2.2-3.3) | 3.7<br>(1.5-6.2) | 5.4<br>(2.6-8.8) | 3.0<br>(1.7-4.0) | 2.8<br>(1.2-4.7) | 1.2<br>(0.2-2.4) | 1.3<br>(0.6-2.1) |
| Non-Hodgkin<br>Lymphoma | 17,940 | 22.5<br>(22.1-22.8) | 23.3<br>(22.9-23.7) | 22.8<br>(19.8-25.8) | 15.6<br>(13.9-17.0) | 11.6<br>(6.9-16.0) | 18.4<br>(11.9-25.2) | 18.5<br>(15.2-22.1) | 14.1<br>(9.9-18.7) | 16<br>(14.2-17.7) | 16<br>(10.0-21.6) | 30.9<br>(24.5-38.6) | 20.9<br>(17.7-23.7) | 18.9<br>(13.1-23.7) | 14.4<br>(8.5-19.3) | 12.6<br>(6.3-19.3) |
| Multiple<br>myeloma | 5,745 | 7.1<br>(6.9-7.3) | 7.1<br>(6.9-7.4) | 7.8<br>(6.2-9.4) | 3.0<br>(2.4-3.7) | 1.0<br>(0.2-2.0) | 2.9<br>(1.0-5.1) | 5.8<br>(4.2-7.8) | 4.1<br>(2.4-6.2) | 7.8<br>(6.5-9.0) | 4.6<br>(2.1-7.4) | 15.3<br>(10.1-21.0) | 14.1<br>(11.8-16.7) | 5.8<br>(3.2-8.9) | 9.0<br>(4.2-14.0) | 17.5<br>(8.2-27.4) |
| Leukemia | 12,010 | 15.4<br>(15.1-15.8) | 16.3<br>(16.0-16.7) | 11.2<br>(9.5-13.0) | 8.6<br>(7.4-9.8) | 2.7<br>(1.2-4.9) | 7.3<br>(4.2-11.4) | 8.2<br>(5.7-10.8) | 7.5<br>(4.9-10.3) | 11.1<br>(9.6-12.6) | 13.8<br>(9.2-19.0) | 12.7<br>(9.0-17.0) | 11.2<br>(8.5-13.5) | 10.8<br>(8.0-14.3) | 7.2<br>(4.4-10.8) | 9.3<br>(3.8-15.2) |

Source: Adapted from Statistics Canada, Canadian Census Health and Environment Cohorts 2006 & 2011, 2006 long-form census, 2011 National Household Survey, Canadian Vital Statistics Death Database 2006-2015, and Canadian Cancer Registry 2006-2015.
95% CI=confidence intervals based on 2.5<sup>th</sup> and 97.5<sup>th</sup> bootstrap percentiles; CNS=central nervous system.
<sup>a</sup> Counts are rounded to base 5.
<sup>b</sup> Population in this group differs by cancer site; includes individuals not classifiable into other racial categories as described in methods, as well as groups with too few cancer cases to meet disclosure threshold.
<sup>c</sup> Number of cases under disclosure threshold, population group was included in 'Other visible minority groups' for this cancer site.

**Table 4.**
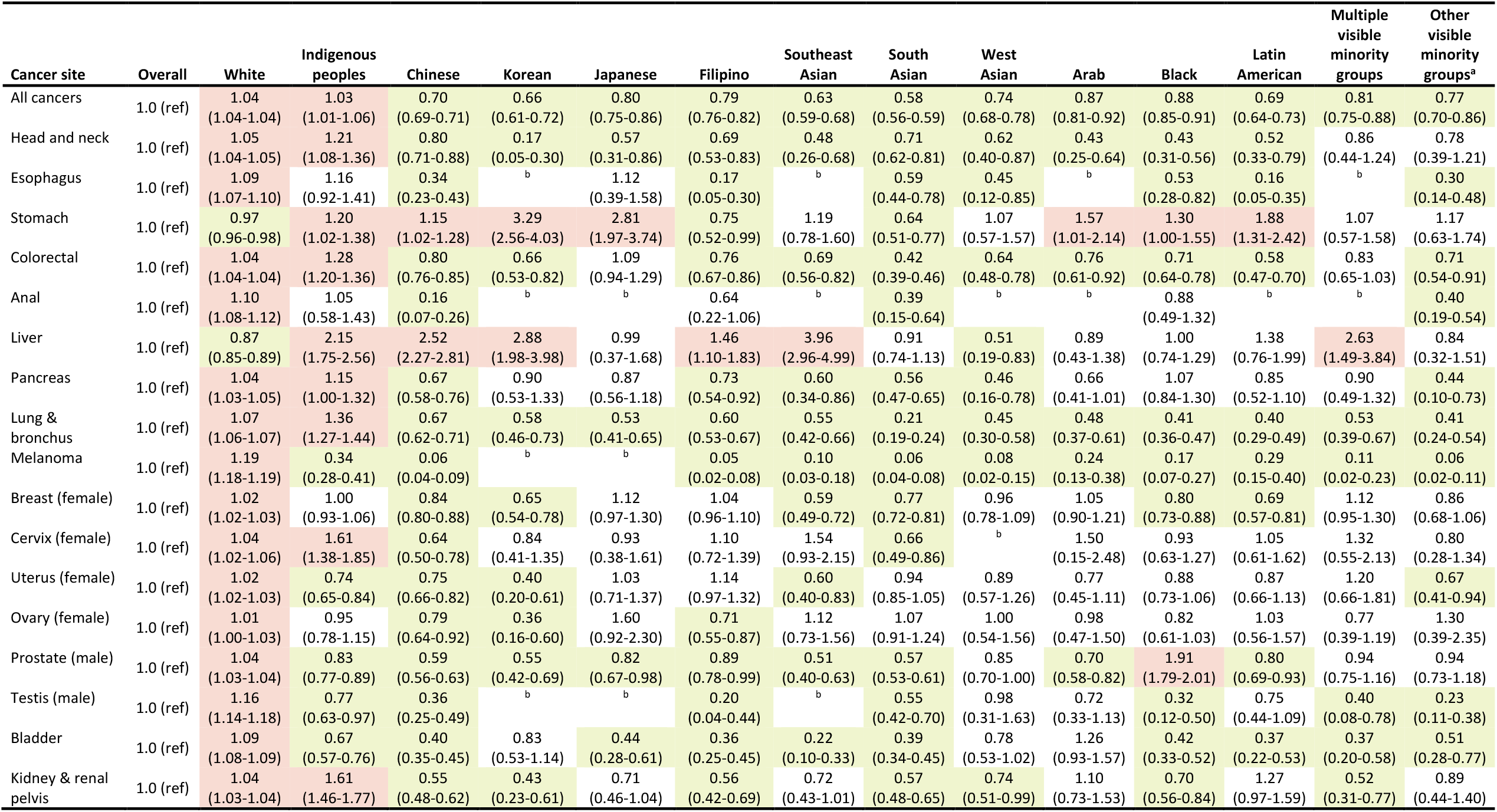
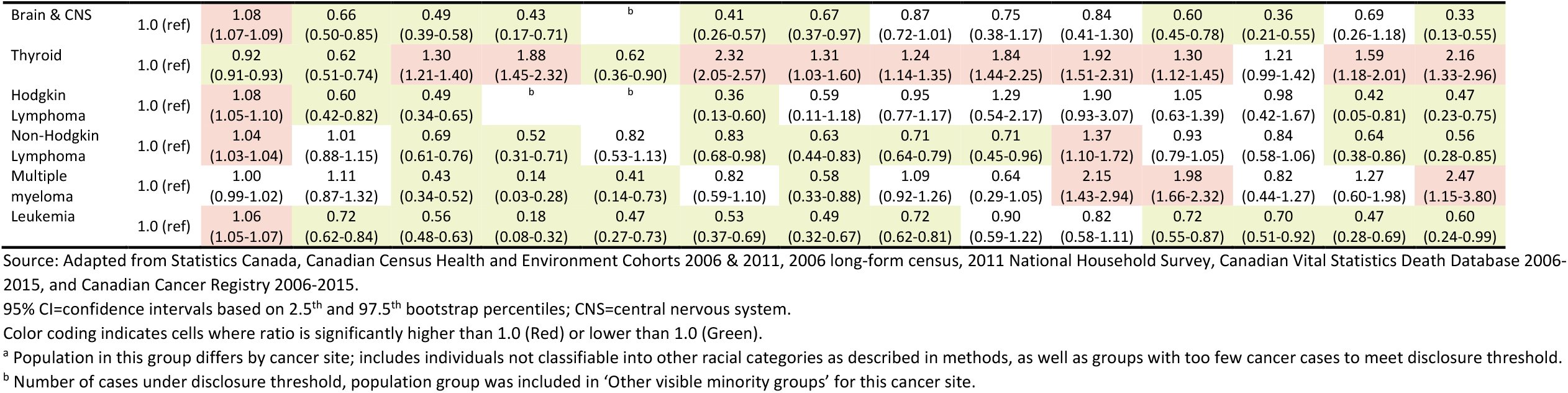
Age-standardized incidence rate ratios by cancer site and by race, age-standardized to the Canadian 2011 census population.

| Cancer site | Overall | White | Indigenous peoples | Chinese | Korean | Japanese | Filipino | Southeast Asian | South Asian | West Asian | Arab | Black | Latin American | Multiple visible minority groups | Other visible minority groups<sup>a</sup> |
| --- | --- | --- | --- | --- | --- | --- | --- | --- | --- | --- | --- | --- | --- | --- | --- |
| All cancers | 1.0 (ref) | 1.04<br>(1.04-1.04) | 1.03<br>(1.01-1.06) | 0.70<br>(0.69-0.71) | 0.66<br>(0.61-0.72) | 0.80<br>(0.75-0.86) | 0.79<br>(0.76-0.82) | 0.63<br>(0.59-0.68) | 0.58<br>(0.56-0.59) | 0.74<br>(0.68-0.78) | 0.87<br>(0.81-0.92) | 0.88<br>(0.85-0.91) | 0.69<br>(0.64-0.73) | 0.81<br>(0.75-0.88) | 0.77<br>(0.70-0.86) |
| Head and neck | 1.0 (ref) | 1.05<br>(1.04-1.05) | 1.21<br>(1.08-1.36) | 0.80<br>(0.71-0.88) | 0.17<br>(0.05-0.30) | 0.57<br>(0.31-0.86) | 0.69<br>(0.53-0.83) | 0.48<br>(0.26-0.68) | 0.71<br>(0.62-0.81) | 0.62<br>(0.40-0.87) | 0.43<br>(0.25-0.64) | 0.43<br>(0.31-0.56) | 0.52<br>(0.33-0.79) | 0.86<br>(0.44-1.24) | 0.78<br>(0.39-1.21) |
| Esophagus | 1.0 (ref) | 1.09<br>(1.07-1.10) | 1.16<br>(0.92-1.41) | 0.34<br>(0.23-0.43) | <sup>b</sup> | 1.12<br>(0.39-1.58) | 0.17<br>(0.05-0.30) | <sup>b</sup> | 0.59<br>(0.44-0.78) | 0.45<br>(0.12-0.85) | <sup>b</sup> | 0.53<br>(0.28-0.82) | 0.16<br>(0.05-0.35) | <sup>b</sup> | 0.30<br>(0.14-0.48) |
| Stomach | 1.0 (ref) | 0.97<br>(0.96-0.98) | 1.20<br>(1.02-1.38) | 1.15<br>(1.02-1.28) | 3.29<br>(2.56-4.03) | 2.81<br>(1.97-3.74) | 0.75<br>(0.52-0.99) | 1.19<br>(0.78-1.60) | 0.64<br>(0.51-0.77) | 1.07<br>(0.57-1.57) | 1.57<br>(1.01-2.14) | 1.30<br>(1.00-1.55) | 1.88<br>(1.31-2.42) | 1.07<br>(0.57-1.58) | 1.17<br>(0.63-1.74) |
| Colorectal | 1.0 (ref) | 1.04<br>(1.04-1.04) | 1.28<br>(1.20-1.36) | 0.80<br>(0.76-0.85) | 0.66<br>(0.53-0.82) | 1.09<br>(0.94-1.29) | 0.76<br>(0.67-0.86) | 0.69<br>(0.56-0.82) | 0.42<br>(0.39-0.46) | 0.64<br>(0.48-0.78) | 0.76<br>(0.61-0.92) | 0.71<br>(0.64-0.78) | 0.58<br>(0.47-0.70) | 0.83<br>(0.65-1.03) | 0.71<br>(0.54-0.91) |
| Anal | 1.0 (ref) | 1.10<br>(1.08-1.12) | 1.05<br>(0.58-1.43) | 0.16<br>(0.07-0.26) | <sup>b</sup> | <sup>b</sup> | 0.64<br>(0.22-1.06) | <sup>b</sup> | 0.39<br>(0.15-0.64) | <sup>b</sup> | <sup>b</sup> | 0.88<br>(0.49-1.32) | <sup>b</sup> | <sup>b</sup> | 0.40<br>(0.19-0.54) |
| Liver | 1.0 (ref) | 0.87<br>(0.85-0.89) | 2.15<br>(1.75-2.56) | 2.52<br>(2.27-2.81) | 2.88<br>(1.98-3.98) | 0.99<br>(0.37-1.68) | 1.46<br>(1.10-1.83) | 3.96<br>(2.96-4.99) | 0.91<br>(0.74-1.13) | 0.51<br>(0.19-0.83) | 0.89<br>(0.43-1.38) | 1.00<br>(0.74-1.29) | 1.38<br>(0.76-1.99) | 2.63<br>(1.49-3.84) | 0.84<br>(0.32-1.51) |
| Pancreas | 1.0 (ref) | 1.04<br>(1.03-1.05) | 1.15<br>(1.00-1.32) | 0.67<br>(0.58-0.76) | 0.90<br>(0.53-1.33) | 0.87<br>(0.56-1.18) | 0.73<br>(0.54-0.92) | 0.60<br>(0.34-0.86) | 0.56<br>(0.47-0.65) | 0.46<br>(0.16-0.78) | 0.66<br>(0.41-1.01) | 1.07<br>(0.84-1.30) | 0.85<br>(0.52-1.10) | 0.90<br>(0.49-1.32) | 0.44<br>(0.10-0.73) |
| Lung & bronchus | 1.0 (ref) | 1.07<br>(1.06-1.07) | 1.36<br>(1.27-1.44) | 0.67<br>(0.62-0.71) | 0.58<br>(0.46-0.73) | 0.53<br>(0.41-0.65) | 0.60<br>(0.53-0.67) | 0.55<br>(0.42-0.66) | 0.21<br>(0.19-0.24) | 0.45<br>(0.30-0.58) | 0.48<br>(0.37-0.61) | 0.41<br>(0.36-0.47) | 0.40<br>(0.29-0.49) | 0.53<br>(0.39-0.67) | 0.41<br>(0.24-0.54) |
| Melanoma | 1.0 (ref) | 1.19<br>(1.18-1.19) | 0.34<br>(0.28-0.41) | 0.06<br>(0.04-0.09) | <sup>b</sup> | <sup>b</sup> | 0.05<br>(0.02-0.08) | 0.10<br>(0.03-0.18) | 0.06<br>(0.04-0.08) | 0.08<br>(0.02-0.15) | 0.24<br>(0.13-0.38) | 0.17<br>(0.07-0.27) | 0.29<br>(0.15-0.40) | 0.11<br>(0.02-0.23) | 0.06<br>(0.02-0.11) |
| Breast (female) | 1.0 (ref) | 1.02<br>(1.02-1.03) | 1.00<br>(0.93-1.06) | 0.84<br>(0.80-0.88) | 0.65<br>(0.54-0.78) | 1.12<br>(0.97-1.30) | 1.04<br>(0.96-1.10) | 0.59<br>(0.49-0.72) | 0.77<br>(0.72-0.81) | 0.96<br>(0.78-1.09) | 1.05<br>(0.90-1.21) | 0.80<br>(0.73-0.88) | 0.69<br>(0.57-0.81) | 1.12<br>(0.95-1.30) | 0.86<br>(0.68-1.06) |
| Cervix (female) | 1.0 (ref) | 1.04<br>(1.02-1.06) | 1.61<br>(1.38-1.85) | 0.64<br>(0.50-0.78) | 0.84<br>(0.41-1.35) | 0.93<br>(0.38-1.61) | 1.10<br>(0.72-1.39) | 1.54<br>(0.93-2.15) | 0.66<br>(0.49-0.86) | <sup>b</sup> | 1.50<br>(0.15-2.48) | 0.93<br>(0.63-1.27) | 1.05<br>(0.61-1.62) | 1.32<br>(0.55-2.13) | 0.80<br>(0.28-1.34) |
| Uterus (female) | 1.0 (ref) | 1.02<br>(1.02-1.03) | 0.74<br>(0.65-0.84) | 0.75<br>(0.66-0.82) | 0.40<br>(0.20-0.61) | 1.03<br>(0.71-1.37) | 1.14<br>(0.97-1.32) | 0.60<br>(0.40-0.83) | 0.94<br>(0.85-1.05) | 0.89<br>(0.57-1.26) | 0.77<br>(0.45-1.11) | 0.88<br>(0.73-1.06) | 0.87<br>(0.66-1.13) | 1.20<br>(0.66-1.81) | 0.67<br>(0.41-0.94) |
| Ovary (female) | 1.0 (ref) | 1.01<br>(1.00-1.03) | 0.95<br>(0.78-1.15) | 0.79<br>(0.64-0.92) | 0.36<br>(0.16-0.60) | 1.60<br>(0.92-2.30) | 0.71<br>(0.55-0.87) | 1.12<br>(0.73-1.56) | 1.07<br>(0.91-1.24) | 1.00<br>(0.54-1.56) | 0.98<br>(0.47-1.50) | 0.82<br>(0.61-1.03) | 1.03<br>(0.56-1.57) | 0.77<br>(0.39-1.19) | 1.30<br>(0.39-2.35) |
| Prostate (male) | 1.0 (ref) | 1.04<br>(1.03-1.04) | 0.83<br>(0.77-0.89) | 0.59<br>(0.56-0.63) | 0.55<br>(0.42-0.69) | 0.82<br>(0.67-0.98) | 0.89<br>(0.78-0.99) | 0.51<br>(0.40-0.63) | 0.57<br>(0.53-0.61) | 0.85<br>(0.70-1.00) | 0.70<br>(0.58-0.82) | 1.91<br>(1.79-2.01) | 0.80<br>(0.69-0.93) | 0.94<br>(0.75-1.16) | 0.94<br>(0.73-1.18) |
| Testis (male) | 1.0 (ref) | 1.16<br>(1.14-1.18) | 0.77<br>(0.63-0.97) | 0.36<br>(0.25-0.49) | <sup>b</sup> | <sup>b</sup> | 0.20<br>(0.04-0.44) | <sup>b</sup> | 0.55<br>(0.42-0.70) | 0.98<br>(0.31-1.63) | 0.72<br>(0.33-1.13) | 0.32<br>(0.12-0.50) | 0.75<br>(0.44-1.09) | 0.40<br>(0.08-0.78) | 0.23<br>(0.11-0.38) |
| Bladder | 1.0 (ref) | 1.09<br>(1.08-1.09) | 0.67<br>(0.57-0.76) | 0.40<br>(0.35-0.45) | 0.83<br>(0.53-1.14) | 0.44<br>(0.28-0.61) | 0.36<br>(0.25-0.45) | 0.22<br>(0.10-0.33) | 0.39<br>(0.34-0.45) | 0.78<br>(0.53-1.02) | 1.26<br>(0.93-1.57) | 0.42<br>(0.33-0.52) | 0.37<br>(0.22-0.53) | 0.37<br>(0.20-0.58) | 0.51<br>(0.28-0.77) |
| Kidney & renal pelvis | 1.0 (ref) | 1.04<br>(1.03-1.04) | 1.61<br>(1.46-1.77) | 0.55<br>(0.48-0.62) | 0.43<br>(0.23-0.61) | 0.71<br>(0.46-1.04) | 0.56<br>(0.42-0.69) | 0.72<br>(0.43-1.01) | 0.57<br>(0.48-0.65) | 0.74<br>(0.51-0.99) | 1.10<br>(0.73-1.53) | 0.70<br>(0.56-0.84) | 1.27<br>(0.97-1.59) | 0.52<br>(0.31-0.77) | 0.89<br>(0.44-1.40) |
| Brain & CNS | 1.0 (ref) | 1.08<br>(1.07-1.09) | 0.66<br>(0.50-0.85) | 0.49<br>(0.39-0.58) | 0.43<br>(0.17-0.71) | <sup>b</sup> | 0.41<br>(0.26-0.57) | 0.67<br>(0.37-0.97) | 0.87<br>(0.72-1.01) | 0.75<br>(0.38-1.17) | 0.84<br>(0.41-1.30) | 0.60<br>(0.45-0.78) | 0.36<br>(0.21-0.55) | 0.69<br>(0.26-1.18) | 0.33<br>(0.13-0.55) |
| Thyroid | 1.0 (ref) | 0.92<br>(0.91-0.93) | 0.62<br>(0.51-0.74) | 1.30<br>(1.21-1.40) | 1.88<br>(1.45-2.32) | 0.62<br>(0.36-0.90) | 2.32<br>(2.05-2.57) | 1.31<br>(1.03-1.60) | 1.24<br>(1.14-1.35) | 1.84<br>(1.44-2.25) | 1.92<br>(1.51-2.31) | 1.30<br>(1.12-1.45) | 1.21<br>(0.99-1.42) | 1.59<br>(1.18-2.01) | 2.16<br>(1.33-2.96) |
| Hodgkin Lymphoma | 1.0 (ref) | 1.08<br>(1.05-1.10) | 0.60<br>(0.42-0.82) | 0.49<br>(0.34-0.65) | <sup>b</sup> | <sup>b</sup> | 0.36<br>(0.13-0.60) | 0.59<br>(0.11-1.18) | 0.95<br>(0.77-1.17) | 1.29<br>(0.54-2.17) | 1.90<br>(0.93-3.07) | 1.05<br>(0.63-1.39) | 0.98<br>(0.42-1.67) | 0.42<br>(0.05-0.81) | 0.47<br>(0.23-0.75) |
| Non-Hodgkin Lymphoma | 1.0 (ref) | 1.04<br>(1.03-1.04) | 1.01<br>(0.88-1.15) | 0.69<br>(0.61-0.76) | 0.52<br>(0.31-0.71) | 0.82<br>(0.53-1.13) | 0.83<br>(0.68-0.98) | 0.63<br>(0.44-0.83) | 0.71<br>(0.64-0.79) | 0.71<br>(0.45-0.96) | 1.37<br>(1.10-1.72) | 0.93<br>(0.79-1.05) | 0.84<br>(0.58-1.06) | 0.64<br>(0.38-0.86) | 0.56<br>(0.28-0.85) |
| Multiple myeloma | 1.0 (ref) | 1.00<br>(0.99-1.02) | 1.11<br>(0.87-1.32) | 0.43<br>(0.34-0.52) | 0.14<br>(0.03-0.28) | 0.41<br>(0.14-0.73) | 0.82<br>(0.59-1.10) | 0.58<br>(0.33-0.88) | 1.09<br>(0.92-1.26) | 0.64<br>(0.29-1.05) | 2.15<br>(1.43-2.94) | 1.98<br>(1.66-2.32) | 0.82<br>(0.44-1.27) | 1.27<br>(0.60-1.98) | 2.47<br>(1.15-3.80) |
| Leukemia | 1.0 (ref) | 1.06<br>(1.05-1.07) | 0.72<br>(0.62-0.84) | 0.56<br>(0.48-0.63) | 0.18<br>(0.08-0.32) | 0.47<br>(0.27-0.73) | 0.53<br>(0.37-0.69) | 0.49<br>(0.32-0.67) | 0.72<br>(0.62-0.81) | 0.90<br>(0.59-1.22) | 0.82<br>(0.58-1.11) | 0.72<br>(0.55-0.87) | 0.70<br>(0.51-0.92) | 0.47<br>(0.28-0.69) | 0.60<br>(0.24-0.99) |

Source: Adapted from Statistics Canada, Canadian Census Health and Environment Cohorts 2006 & 2011, 2006 long-form census, 2011 National Household Survey, Canadian Vital Statistics Death Database 2006-2015, and Canadian Cancer Registry 2006-2015.
95% CI=confidence intervals based on 2.5<sup>th</sup> and 97.5<sup>th</sup> bootstrap percentiles; CNS=central nervous system.
Color coding indicates cells where ratio is significantly higher than 1.0 (Red) or lower than 1.0 (Green).
<sup>a</sup> Population in this group differs by cancer site; includes individuals not classifiable into other racial categories as described in methods, as well as groups with too few cancer cases to meet disclosure threshold.
<sup>b</sup> Number of cases under disclosure threshold, population group was included in 'Other visible minority groups' for this cancer site.

Differences in most cancer incidence rates between racial groups persisted even after adjustment for age, sex, year, province, place of residence, education level, and household income in regression models (Table 5). Stratification by immigration status further revealed relationships with immigration which differed by site. For most cancer sites, the cancer incidence rates showed a trend of convergence, where native-born members of racial groups had closer incidence rates to the native-born White individuals than foreign-born members of racial groups. This was not the case for all cancer sites, however: stomach, cervical, thyroid cancer, and multiple myeloma incidence rates were in most cases equivalent between foreign-born and native-born racial groups.

**Table 5.**
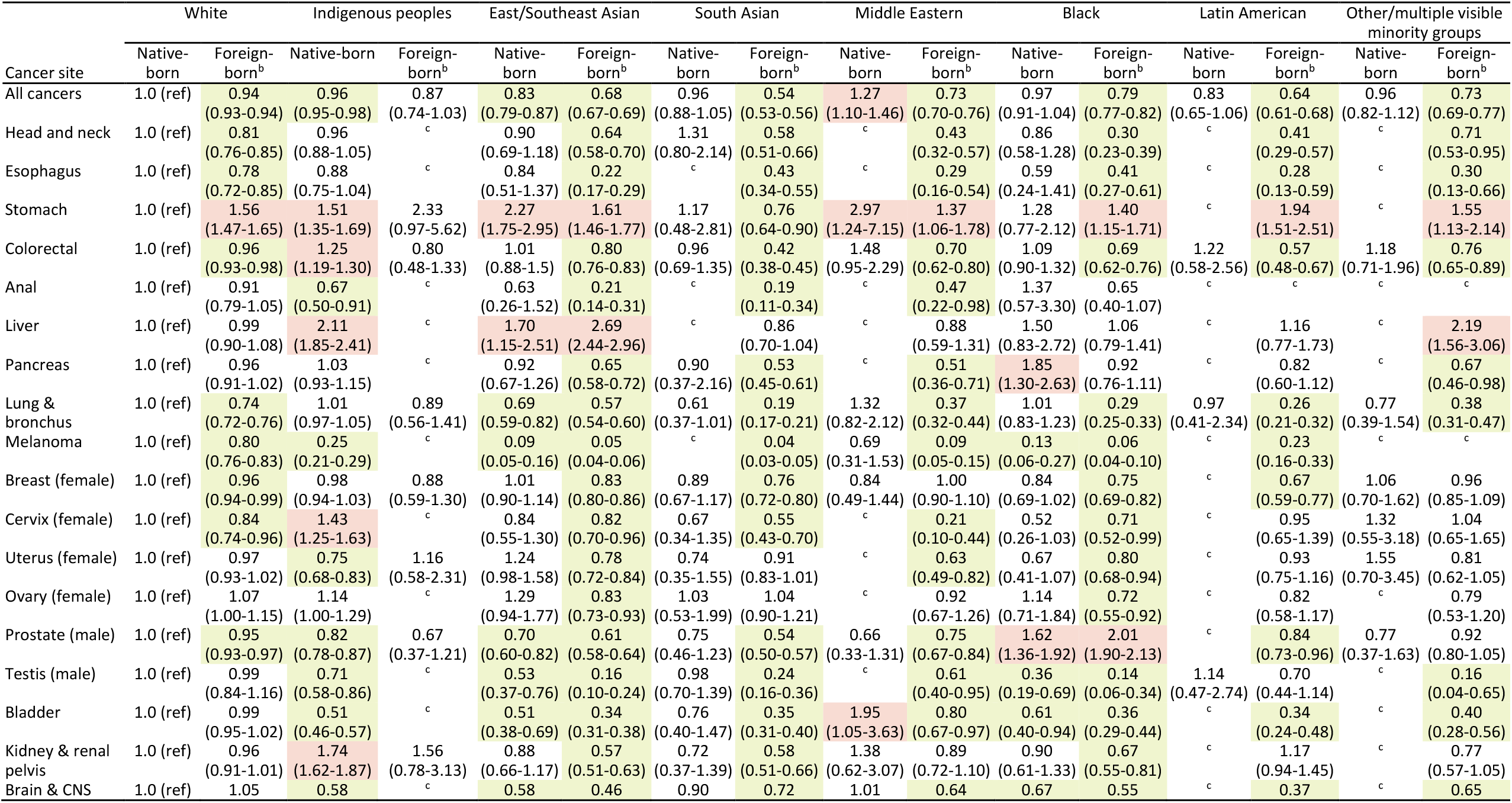
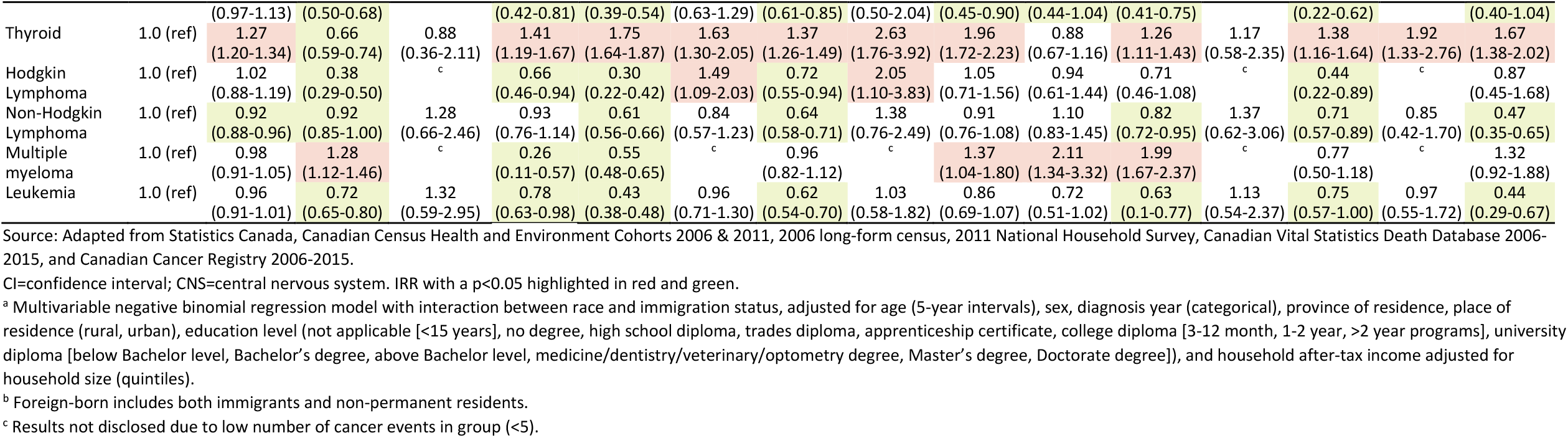
Negative binomial regression cancer incidence rate ratios by race and immigration status, adjusted for age, sex, year, province, and socioeconomic variables.^a^.

Age-standardized incidence rates by visible minority groups compared with GLOBOCAN 2020 incidence estimates by world regions of origin are provided in Supplementary Table 4 and Supplementary Figures 1-24. For most cancer sites, incidence rates tended to be intermediate between the rates in White Canadians and the rates from their world region of origin. However, for head and neck, lung and bronchus, esophageal, pancreatic, and cervical cancers, the incidence rates in visible minority groups were markedly lower than in both White Canadians and their world region of origin. For thyroid cancer, the incidence rates in visible minority groups were strikingly higher than in both White Canadians and their world region of origin.

## Discussion

This study estimated cancer incidence rates by race using a very large representative population-based sample of the Canadian population linked to a cancer registry with national coverage. Because the CCR does not collect data on race, our results are to our knowledge the most nationally representative up-to-date estimates of cancer incidence rates stratified by race in Canada. We found that for most cancer sites, non-White and non-Indigenous racial groups had lower age-standardized cancer incidence rates than the overall population. Indigenous peoples had higher age-standardized incidence rates for some cancer sites but lower for others compared with the overall population. Differences in cancer rates between racial groups remained even after adjusting for differences in socioeconomic status. However, immigration status was an important modifier of cancer risk, suggesting that a healthy immigrant effect influences cancer risk by race in Canada.

Inequalities in cancer incidence are influenced by the social determinants of health.^9,27^ Individuals who are more resourced are in general better able to adopt healthy lifestyles and avail themselves of preventive healthcare.^28^ Though Canada has a public healthcare system with universal health coverage available for all citizens and permanent residents, there is a socioeconomic gradient in cancer risk across multiple sites.^29-32^ Nevertheless, our analysis suggests that differences in household income, education, and urbanicity are not the fundamental causes of the observed differences in cancer incidence observed between racial groups, as incidence differences persisted despite adjustment for these variables.

Systemic racism and enduring colonial influences on Indigenous peoples’ health contribute to health inequalities between racial groups.^33^ However, the observed patterns suggest that differences between racial groups are also likely linked to lifestyles, culture, early life exposures, and selection factors for immigration.

Immigration was an important modifier of cancer risk. Most immigrants to Canada are economic immigrants selected based on their ability to contribute to labour market needs.^34^ Immigrants applying for permanent residency must undergo a medical examination. Applicants may be found inadmissible for residency if they present medical conditions deemed to potentially cause excessive demands on health and social services. There is therefore a strong selection for immigrants in better health than native-born Canadians, known as the healthy immigrant effect.^35,36^ We hypothesize that cancer rates would be intermediate between the world region of origin and those in native-born Canadians for cancer sites where adult lifestyles are important risk factors such as obesity, diet, and occupational exposures. We found that this was the case for many cancer sites such as colorectal, breast, uterus (corpus), prostate, kidney and renal pelvis, non-Hodgkin lymphoma, and leukemia. However, for many other sites the cancer incidence rate in racial groups was lower than in both native-born Canadians and their world region of origin. This was the case for head and neck, esophagus, lung and bronchus, and cervical cancers. These cancers are strongly related to exposures adopted early in life prior to immigration and linked with socioeconomic status: tobacco consumption, alcohol drinking, and early life infections.^37,38^ The low incidence rates for these cancers suggest there is a selection for immigrants in better health and from higher socioeconomic backgrounds from within their country of origin, who are less likely to be smokers^36^ and heavy drinkers,^39^ and exposed to some infections. The high rates of cervical cancer in Indigenous women are attributable to both higher human papillomavirus (HPV) infection prevalence and lower screening coverage;^40-43^ however we are unaware of any studies comparing HPV prevalence by immigrant status or other racial groups in Canada. Immigrant and racialized women are nonetheless known to have lower cervical cancer screening rates,^5,44^ suggesting that the lower risk of cervical cancer in many visible minority groups cannot be attributed to more screening and is likely in part due to a lower prevalence of HPV. Lastly, for some cancer sites the cancer incidence rates were higher in some racial groups than in both White native-born Canadians and their world region of origin, notably for prostate (Black men) and thyroid cancers, and for multiple myeloma (some groups). As these cancers are often diagnosed through incidental findings, this pattern potentially signals underdiagnosis of these cancers in some world regions, and more incidental diagnoses in Canada during examinations for other health problems.

Stomach, liver, and thyroid stood out as the cancer sites where incidence rates were systematically higher across most racial groups than in White individuals. It is likely that for stomach and liver cancers, the higher incidence rates reflect childhood exposures to *Helicobacter pylori* and hepatitis B virus in the countries of origin of many immigrants,^45,46^ as well as higher prevalences of *H. pylori* and hepatitis C virus in some Indigenous populations.^46-48^ Remarkably, the rates of these cancers were also high in Canadian-born East Asian, Southeast Asian, and Middle Eastern individuals. It is possible that the persisting high rates of these cancers in native-born Canadians of these racial groups are due to vertical or familial transmission of these pathogens. While all provinces perform prenatal screening for hepatitis B to provide vaccination against hepatitis B to at-risk children, prenatal hepatitis B screening and follow-up has imperfect coverage.^49^ There is no routine screening for *H. pylori* in Canada. The risk of stomach cancer does not appear to decline with time since immigration in many Canadian immigrant groups.^50^ While *H. pylori* prevalence tends to decrease in successive generations of immigrants, it still often remains higher than the prevalence of the native host country population.^51^ Persistence of stomach cancer risk across generations may also reflect conserved cultural differences in diet, such as consumption of foods high in salt.^52^ For thyroid cancer, more incidental cancer diagnoses in the Canadian healthcare system likely account for the higher rates of this cancer in visible minority groups compared with their world regions of origin. However, it is not clear why rates are higher in visible minority groups than in White Canadians and Indigenous peoples. This finding is in stark contrast with the United States, where thyroid cancer incidence rates are highest in White populations and lowest in Black populations due to racial disparities in access to healthcare.^53^ Non-White racial groups are less likely to use cancer screening services and just as likely to have contact with specialist physicians as White Canadians,^54^ so the higher thyroid cancer rates are not plausibly explained by more incidental findings in visible minority groups.

An important limitation of our analysis is that we are unable to account for emigration; it is therefore possible we may have underestimated cancer incidence rates due to the inclusion of person-time from cohort members who have left Canada. Emigration is likely higher for non-permanent residents. Cancer incidence might therefore be more underestimated in groups with more non-permanent residents, such as Korean and Latin American populations. Cancer incidence may also be underestimated due to the exclusion from CanCHECs of homeless or institutionalized populations living in nursing homes and penitentiaries, who are likely to be in poorer health. However, we compared CanCHECs cancer incidence rates with the national incidence rates (which are not subject to these biases) and found very similar rates. This suggests that this underestimation is likely to be minimal overall. We aimed to provide as disaggregated data as possible for racial categories, in alignment with calls for disaggregated health data by race.^1,8^ However, this disaggregation came at the cost of lower statistical power for some cancer sites to detect meaningful differences. Even by pooling two CanCHECs with several years of follow-up, we were unable to meet confidentiality disclosure thresholds for rarer cancers in some racial groups.

While previous studies have examined racial and ethnic differences in cancer rates in Canada, they have tended to be based in a single province or examined only the most frequent cancer types. Much previous research has focused on cancer in Indigenous peoples,^6,40,55,56^ with much less research on other racial groups. A study of the 1991 CanCHEC established the existence of a healthy immigrant effect for cancer incidence rates by immigration status.^3^ It is often assumed that cancer rates in immigrants will trend towards those in their host country with time since immigration due to acculturation. However, that study and others^35^ found that the convergence of cancer incidence rates and mortality rates between immigrants and non-immigrants over years since arrival might be partly explained by changes in the countries of origin of immigrants over time, as more recent immigrants have come from world regions with lower cancer rates. These findings suggest that either immigrant lifestyles might not assimilate as much as previously believed, or that lifestyle changes later in adulthood may be less important for cancer risk and point to a potentially important role of early life exposures. For example, the proportion of overweight immigrants does not greatly increase with time since immigration, and remains lower than in Canadian-born individuals even a decade after immigration.^36^ While there are few studies of dietary acculturation in Canada, some studies suggest that many immigrants maintain their traditional diet to a certain extent.^36,57^ If culture and lifestyle are conserved across generations, this may explain why we found that cancer incidence did not fully converge between Canadian-born visible minorities and those not identifying as visible minorities. A previous study of the 2006 CanCHEC estimated differences in overall cancer incidence rates by ethnicity and immigration status,^4^ but did not estimate site-specific cancer incidence rates as we have done. That study also examined the construct of ethnicity rather than race, using the ethnic origin census question rather than the Indigenous group and visible minority group questions. While there is construct overlap, a person’s ethnic origin may not necessarily correspond with their race. We opted to use the constructs of visible minority group combined with Indigenous identity to capture race rather than ethnicity, and to align more closely with race-based and Indigenous identity data standards for measuring racial health inequities in Canada.^11^

Cancer is a heterogenous group of diseases with different aetiologies and long latency. We focus the discussion above on large trends across sites; however, an understanding of the drivers of cancer inequalities requires a separate analysis by cancer site and over time to account for changes in risk after adoption of a new home country. Lastly, the large differences we have observed between racial groups point to important potential gains in cancer prevention. The ComPARe study estimated that 33-37% of cancers among adults in Canada are attributable to preventable risk factors such as tobacco consumption.^38^ However, the differences we observed between racial groups are in some cases larger than the estimated preventable fractions of some cancer sites. This suggests that the preventable fraction of cancer from environment and lifestyle may still be underestimated. More studies of immigrant and racial group lifestyles and experiences may further elucidate opportunities for cancer prevention.

## Data Availability

Statistics Canada is the owner and steward of the data used in this report, and access to the data is regulated by the 1985 Statistics Act. To access the data, researchers must become deemed employees of Statistics Canada and sign a research contract. Members of post-secondary institutions such as a faculty, students, or staff may apply for data access to Statistics Canada microdata through the Research Data Centre program using the Microdata Access Portal (https://www.statcan.gc.ca/en/microdata/data-centres/access). Code used for the current analyses is available at the Borealis repository, at https://doi.org/10.5683/SP3/Z8RBON.

https://www.statcan.gc.ca/en/microdata/data-centres/access

## Conflicts of interest

TM, SM, and PT have no conflicts of interest to declare. ELF reports grants to his institution from the Canadian Institutes of Health Research, the National Institutes of Health, and Merck during the conduct of the study; and personal fees from Merck. MZ and ELF hold a patent related to the discovery “ DNA methylation markers for early detection of cervical cancer”, registered at the Office of Innovation and Partnerships, McGill University, Montréal, Québec, Canada.

## Funding

This study was funded by a Canadian Institutes of Health Research (CIHR) (grant 179901) to EF, and by a CIHR HIV/AIDS and STBBI Research Initiative, sponsored by the CIHR Institute of Infection and Immunity, Postdoctoral Fellowship Award (support to SM; Funding Reference Number: 202110HIV-477526-93701).

## Acknowledgement

Data Source: Statistics Canada, Canadian Census Health and Environment Cohorts 2006 & 2011, 2006 long-form census, 2011 National Household Survey, Canadian Vital Statistics Death Database 2006-2015, and Canadian Cancer Registry 2006-2015. The Postal Code^OM^ Conversion File Plus (7D) is based on data licensed from Canada Post Corporation. Reproduced and distributed on an “ as is” basis with the permission of Statistics Canada. This does not constitute an endorsement by Statistics Canada of this product.

## Data availability

Statistics Canada is the owner and steward of the data used in this report, and access to the data is regulated by the 1985 *Statistics Act*. To access the data, researchers must become deemed employees of Statistics Canada and sign a research contract. Members of post-secondary institutions such as a faculty, students, or staff may apply for data access to Statistics Canada microdata through the Research Data Centre program using the Microdata Access Portal (https://www.statcan.gc.ca/en/microdata/data-centres/access). Code used for the current analyses is available at the Borealis repository, at https://doi.org/10.5683/SP3/Z8RBON.

## Notes

### Competing Interest Statement

TM, SM, and PT have no conflicts of interest to declare. ELF reports grants to his institution from CIHR, the National Institutes of Health, and Merck during the conduct of the study; and personal fees from Merck. MZ and ELF hold a patent related to the discovery "DNA methylation markers for early detection of cervical cancer", registered at the Office of Innovation and Partnerships, McGill University, Montreal, Quebec, Canada.

### Author Declarations

Ethics committee/IRB of McGill University gave ethical approval for this work.

