## Supplementary Appendix for "Site-specific cancer incidence by race and immigration status in Canada 2006-2015: a population-based data linkage study"

### Supplementary Tables

Supplementary Table 1. ICD-O-3 Topography and histology codes by cancer site based on SEER site groupings.

| Cancer site | ICD-O-3 Topography | ICD-O-3 Histology (Type) |
|---|---|---|
| All cancers | All sites C00-C80 | All invasive sites |
| Head and neck[a] | C00-C14, C300-C329 | 8000-9049, 9056-9139,9141-9589 |
| Esophagus | C150-C159 | 8000-9049, 9056-9139,9141-9589 |
| Stomach | C160-C169 | 8000-9049, 9056-9139,9141-9589 |
| Colorectal | C180-C189, C260, C199, C209 | 8000-9049, 9056-9139,9141-9589 |
| Anal | C210-C212, C218 | 8000-9049, 9056-9139,9141-9589 |
| Liver | C220 | 8000-9049, 9056-9139,9141-9589 |
| Pancreas | C250-C259 | 8000-9049, 9056-9139,9141-9589 |
| Lung & bronchus | C340-C349 | 8000-9049, 9056-9139,9141-9589 |
| Melanoma | C440-C449 | 8720-8790 |
| Breast | C500-C509 | 8000-9049, 9056-9139,9141-9589 |
| Cervix (female) | C530-C539 | 8000-9049, 9056-9139,9141-9589 |
| Uterus (female) | C540-C549, C559 | 8000-9049, 9056-9139,9141-9589 |
| Ovary (female) | C569 | 8000-9049, 9056-9139,9141-9589 |
| Prostate (male) | C619 | 8000-9049, 9056-9139,9141-9589 |
| Testis (male) | C620-C629 | 8000-9049, 9056-9139,9141-9589 |
| Bladder | C670-C679 | 8000-9049, 9056-9139,9141-9589 |
| Kidney & renal pelvis | C649, C659 | 8000-9049, 9056-9139,9141-9589 |
| Brain & CNS | C710-C719 | 8000-9049, 9056-9139,9141-9589 |
| | C710-C719 | 9530 - 9539 |
| | C700-C709, C720-C729 | 8000-9049, 9056-9139,9141-9589 |
| Thyroid | C739 | 8000-9049, 9056-9139,9141-9589 |
| Hodgkin Lymphoma | C000-C809 | 9650-9667 |
| Non-Hodgkin Lymphoma | C000-C809 | 9590-9597, 9670-9729, 9735-9738 |
| | All topographies excluding (C420, C421, C424) | 9811-9818, 9823, 9827, 9837 |
| Multiple myeloma | C000-C809 | 9731-9732, 9734 |
| Leukemia | C000 - C809 | 9826, 9835-9836 |
| | C420, C421, C424 | 9811-9818, 9837 |
| | C420, C421, C424 | 9840, 9861, 9865, 9866, 9867, 9869, 9871-9874, 9895-9897, 9898, 9910, 9911, 9920 |
| | C000 - C809 | 9863, 9875, 9876, 9945, 9946 |
| | C000 - C809 | 9733, 9742, 9800, 9801, 9805, 9806, 9807, 9808, 9809, 9820, 9831, 9832, 9833, 9834, 9860, 9870, 9891, 9930, 9931, 9940, 9948, 9963, 9964 |
| | C420, C421, C424 | 9827 |

CNS=central nervous system; ICD-O-3= International Classification of Diseases for Oncology, 3rd Edition; SEER=Surveillance, Epidemiology, and End Results.

[a] Definition based on the Canadian Cancer Statistics 2021 rather than SEER site groupings.

Supplementary Table 2. Comparison of age-standardized cancer incidence rates per 100,000 person-years by cancer site and by sex between CanCHECs and the Canadian Cancer Registry for Canada, excluding Québec, 2013.

| | Both sexes | | Males | | Females | |
|---|---|---|---|---|---|---|
| Cancer site | CanCHECs 2006 & 2011 (95%CI) | Canadian Cancer Registry (2013) | CanCHECs 2006 & 2011 (95%CI) | Canadian Cancer Registry (2013) | CanCHECs 2006 & 2011 (95%CI) | Canadian Cancer Registry (2013) |
| All cancers | 514.0 (512.1-515.7) | 524.3 | 570.2 (567.5-573.0) | 577.9 | 471.3 (468.8-474.0) | 486.4 |
| Head and neck | 15.4 (15.0-15.7) | 15.7 | 23.2 (22.5-23.8) | 23.9 | 8.4 (8.0-8.7) | 8.2 |
| Esophagus | 5.2 (5.0-5.4) | 5.7 | 8.4 (8.2-9.0) | 9.3 | 2.2 (2.1-2.4) | 2.5 |
| Stomach | 9.3 (9.0-9.5) | 9.3 | 13.3 (12.8-13.7) | 13.1 | 6.0 (5.7-6.3) | 6.2 |
| Colorectal | 62.4 (61.8-63.0) | 62.2 | 74.6 (73.6-75.6) | 74.5 | 51.9 (51.0-52.6) | 51.6 |
| Anal | 1.8 (1.7-2.0) | 1.8 | 1.4 (1.3-1.5) | 1.2 | 2.2 (2.1-2.4) | 2.3 |
| Liver | 5.1 (4.9-5.3) | 6.5 | 8.1 (7.8-8.5) | 10.3 | 2.4 (2.2-2.6) | 3.1 |
| Pancreas | 12.4 (12.1-12.7) | 14.3 | 13.9 (13.4-14.3) | 16.1 | 11.2 (10.8-11.6) | 12.7 |
| Lung & bronchus | 66.0 (65.3-66.7) | 66.9 | 74.3 (73.2-75.3) | 74.6 | 59.9 (59.0-60.7) | 61.4 |
| Melanoma | 19.3 (19.0-19.7) | 20.6 | 22.3 (21.9-22.9) | 23.5 | 17.0 (16.6-17.5) | 18.5 |
| Breast | 67.3 (66.6-68.0) | 66.5 | 1.1 (1.0-1.2) | 1.1 | 127.1 (125.8-128.4) | 126.6 |
| Cervix (female) | - | - | - | - | 7.4 (7.1-7.7) | 7.7 |
| Uterus (female) | - | - | - | - | 31.0 (30.4-31.7) | 31.5 |
| Ovary (female) | - | - | - | - | 14.2 (13.7-14.6) | 15.3 |
| Prostate (male) | - | - | 138.5 (137.3-140.0) | 123.3 | - | - |
| Testis (male) | - | - | 5.9 (5.6-6.2) | 5.8 | - | - |
| Bladder | 24.0 (23.6-24.4) | 25.7 | 40.1 (39.5-41.0) | 43.7 | 10.8 (10.4-11.2) | 11.1 |
| Kidney & renal pelvis | 15.4 (15.0-15.7) | 16.0 | 20.4 (19.9-21.0) | 21.6 | 10.9 (10.5-11.2) | 11.0 |
| Brain & CNS | 7.5 (7.3-7.7) | 7.7 | 8.6 (8.3-9.0) | 9.1 | 6.5 (6.2-6.8) | 6.3 |
| Thyroid | 16.1 (15.8-16.5) | 17.8 | 7.9 (7.6-8.3) | 8.8 | 23.9 (23.2-24.5) | 26.6 |
| Hodgkin Lymphoma | 2.8 (2.7-3.0) | 2.7 | 3.2 (3.0-3.4) | 3.1 | 2.5 (2.3-2.7) | 2.2 |
| Non-Hodgkin Lymphoma | 22.5 (22.1-22.8) | 24.4 | 26.4 (25.6-27.0) | 29.0 | 19.2 (18.7-19.7) | 20.5 |
| Multiple myeloma | 7.1 (6.9-7.3) | 7.7 | 8.5 (8.2-8.9) | 10.0 | 5.9 (5.7-6.2) | 5.9 |
| Leukemia | 15.4 (15.1-15.8) | 16.4 | 19.7 (19.2-20.3) | 21.1 | 11.9 (11.5-12.3) | 12.6 |

Source: Adapted from Statistics Canada, Canadian Census Health and Environment Cohorts 2006 & 2011, 2006 long-form census, 2011 National Household Survey, Canadian Vital Statistics Death Database 2006-2015, and Canadian Cancer Registry 2006-2015. This does not constitute an endorsement by Statistics Canada of this product.

CanCHECs=Canadian Census Health and Environment Cohorts; CI=confidence intervals; CNS=central nervous system.

Rates are age-standardized to the 2011 Canadian census population.

Supplementary Table 3. Comparison of age-specific overall cancer incidence rate (/100,000 person-years) by age group between CanCHECs and the Canadian Cancer Registry for Canada, excluding Québec, 2013.

| | **Age-specific incidence rate (/100,000)** | |
|---|---|---|
| **Age group** | CanCHECs 2006 & 2011 | Canadian Cancer Registry (2013) |
| **0 to 4 years** | 22.8 | 25.7 |
| **5 to 9 years** | 12.6 | 13.2 |
| **10 to 14 years** | 13.9 | 14.8 |
| **15 to 19 years** | 22.9 | 23.1 |
| **20 to 24 years** | 38.4 | 35.1 |
| **25 to 29 years** | 57.3 | 56.9 |
| **30 to 34 years** | 87.4 | 90.9 |
| **35 to 39 years** | 128.5 | 133.8 |
| **40 to 44 years** | 197.9 | 209.0 |
| **45 to 49 years** | 316.2 | 317.7 |
| **50 to 54 years** | 501.7 | 510.4 |
| **55 to 59 years** | 769.0 | 762.1 |
| **60 to 64 years** | 1122.7 | 1111.2 |
| **65 to 69 years** | 1586.1 | 1577.6 |
| **70 to 74 years** | 1976.1 | 2021.9 |
| **75 to 79 years** | 2253.8 | 2313.8 |
| **80 to 84 years** | 2418.2 | 2548.9 |
| **85 to 89 years** | 2469.4 | 2649.7 |
| **90 years and over** | 2121.2 | 2380.0 |

Source: Adapted from Statistics Canada, Canadian Census Health and Environment Cohorts 2006 & 2011, 2006 long-form census, 2011 National Household Survey, Canadian Vital Statistics Death Database 2006-2015, Canadian Cancer Registry 2006-2015. This does not constitute an endorsement by Statistics Canada of this product.

CanCHECs=Canadian Census Health and Environment Cohorts.

Supplementary Table 4. Comparison of age-standardized cancer incidence rates per 100,000 person-years between CanCHECs and GLOBOCAN 2020 world regions of origin, standardized to the world standard population.

| | White | Indigenous peoples | Chinese | | Korean | | Japanese | | Filipino | | Southeast Asian | | South Asian | | West Asian | | Arab | | Black | | Latin American | |
|---|---|---|---|---|---|---|---|---|---|---|---|---|---|---|---|---|---|---|---|---|---|---|
| Cancer site | CanCHECs | CanCHECs | CanCHECs | GLOBOCAN 2020[a] | CanCHECs | GLOBOCAN 2020[b] | CanCHECs | GLOBOCAN 2020[c] | CanCHECs | GLOBOCAN 2020[d] | CanCHECs | GLOBOCAN 2020[e] | CanCHECs | GLOBOCAN 2020[f] | CanCHECs[g] | GLOBOCAN 2020[h] | CanCHECs | GLOBOCAN 2020[i] | CanCHECs | GLOBOCAN 2020[j] | CanCHECs | GLOBOCAN 2020[k] |
| All cancers | 306 | 299 | 208 | 204 | 196 | 240 | 231 | 283 | 243 | 161 | 192 | 150 | 178 | 101 | 237 | 172 | 257 | 157 | 264 | 124 | 208 | 178 |
| Head and neck | 9.3 | 10.9 | 7.7 | 6.6 | 1.9 | 5.5 | 5.3 | 8.0 | 6.3 | 7.7 | 4.4 | 11.1 | 6.1 | 16.2 | 6.2 | 6.5 | 4.0 | 6.3 | 4.2 | 5.0 | 4.8 | 6.7 |
| Esophagus | 3 | 3.2 | 0.9 | 13.8 | [l] | 2.4 | 2.5 | 7.2 | 0.4 | 1.2 | [l] | 2.0 | 1.5 | 5.6 | 1.5 | 1.7 | [l] | 1.6 | 1.6 | 4.4 | 0.5 | 2.4 |
| Stomach | 4.6 | 6.1 | 5.7 | 20.6 | 15.6 | 27.9 | 12.7 | 31.6 | 3.9 | 3.7 | 5.4 | 5.5 | 3.2 | 5.5 | 5.8 | 8.5 | 6.7 | 6.6 | 5.9 | 4.2 | 8.5 | 8.3 |
| Colorectal | 34 | 42.5 | 25.8 | 23.7 | 23.3 | 26.9 | 36.1 | 38.3 | 25.6 | 18.6 | 22.6 | 14.5 | 14.6 | 5.1 | 22.3 | 16.5 | 26.8 | 13.2 | 24.0 | 7.1 | 18.8 | 16.0 |
| Anal | 1.2 | 1.1 | 0.2 | 0.2 | [l] | 0.3 | [l] | 0.3 | 0.8 | 0.2 | [l] | 0.3 | 0.3 | 0.4 | [l] | 0.3 | [l] | 0.3 | 1.0 | 0.9 | [l] | 0.7 |
| Liver | 2.5 | 5.9 | 7.2 | 18.2 | 8.0 | 14.3 | 2.5 | 10.4 | 3.6 | 11.4 | 10.8 | 13.7 | 2.4 | 3.0 | 1.7 | 4.7 | 2.3 | 9.5 | 3.2 | 6.3 | 3.7 | 4.8 |
| Pancreas | 6.6 | 7.6 | 3.9 | 5.3 | 5.7 | 7.1 | 4.7 | 9.9 | 4.4 | 3.7 | 4.4 | 2.3 | 3.6 | 1.2 | 2.9 | 5.7 | 4.4 | 4.4 | 6.8 | 2.1 | 5.6 | 4.5 |
| Lung & bronchus | 36.6 | 46.6 | 21.3 | 34.8 | 17.6 | 25.5 | 15.9 | 32.1 | 20.1 | 21.1 | 18.0 | 17.2 | 7.1 | 6.6 | 13.6 | 24.2 | 17.1 | 18.1 | 14.0 | 4.3 | 13.2 | 12.0 |
| Melanoma | 14.3 | 4.1 | 0.8 | 0.4 | [l] | 0.7 | [l] | 0.5 | 0.7 | 0.5 | 1.0 | 0.5 | 0.7 | 0.3 | 1.5 | 1.5 | 2.0 | 1.0 | 1.7 | 1.1 | 3.5 | 2.3 |
| Breast (female) | 79.8 | 76.1 | 68.9 | 39.1 | 55.6 | 64.2 | 89.9 | 76.3 | 84.6 | 52.7 | 46.7 | 41.2 | 61.1 | 26.2 | 81.3 | 46.6 | 83.0 | 48.0 | 62.7 | 37.8 | 53.9 | 51.9 |
| Cervix (female) | 5.9 | 8.8 | 3.4 | 10.7 | 4.0 | 8.1 | 4.8 | 15.2 | 6.1 | 15.2 | 7.7 | 17.8 | 3.2 | 15.3 | | 4.1 | 3.4 | 5.1 | 4.3 | 32.2 | 5.1 | 14.9 |
| Uterus (female) | 19.1 | 14.5 | 14.5 | 7.6 | 7.9 | 8.1 | 18.9 | 15.0 | 21.7 | 8.7 | 12.1 | 6.6 | 18.0 | 2.7 | 15.0 | 8.8 | 14.0 | 6.3 | 16.1 | 3.5 | 16.7 | 8.2 |
| Ovary (female) | 8.7 | 7.8 | 7.1 | 5.3 | 3.5 | 6.5 | 16.3 | 9.3 | 7.6 | 10.4 | 9.4 | 8.1 | 9.7 | 6.2 | 10.0 | 6.6 | 8.0 | 6.2 | 7.1 | 5.3 | 9.0 | 5.8 |
| Prostate (male) | 78.7 | 60.8 | 43.7 | 10.2 | 43.6 | 27.3 | 59.5 | 51.8 | 64.7 | 23.4 | 40.2 | 13.5 | 42.4 | 6.3 | 68.2 | 28.6 | 51.0 | 22.9 | 153.6 | 35.4 | 62.8 | 59.2 |
| Testis (male) | 6.5 | 4.3 | 2.1 | 0.6 | [l] | 1.4 | [l] | 4.2 | 0.7 | 0.7 | [l] | 0.8 | 3.4 | 0.7 | 5.6 | 1.8 | 4.1 | 1.3 | 1.7 | 0.6 | 4.3 | 3.8 |
| Bladder | 13.1 | 8.1 | 4.6 | 3.6 | 7.5 | 4.3 | 4.5 | 8.2 | 4.1 | 1.9 | 2.8 | 2.6 | 4.8 | 1.9 | 9.1 | 8.6 | 15.6 | 8.8 | 4.8 | 2.7 | 4.7 | 4.0 |
| Kidney & renal pelvis | 9.5 | 14.7 | 5.1 | 3.3 | 4.3 | 6.5 | 5.8 | 7.6 | 5.2 | 2.5 | 7.1 | 1.7 | 5.4 | 1.4 | 7.8 | 4.1 | 9.4 | 3.2 | 6.7 | 1.6 | 11.4 | 4.7 |
| Brain & CNS | 5.9 | 3.8 | 2.6 | 4.1 | 2.3 | 3.0 | [l] | 2.7 | 2.6 | 2.0 | 5.7 | 2.4 | 5.1 | 2.5 | 4.6 | 5.0 | 4.4 | 4.5 | 3.6 | 1.2 | 2.4 | 3.5 |
| Thyroid | 11 | 7.4 | 15.5 | 11.3 | 22.3 | 26.6 | 7.4 | 8.0 | 27.2 | 6.2 | 15.1 | 4.4 | 15.3 | 1.6 | 23.3 | 8.6 | 22.9 | 6.4 | 14.9 | 1.5 | 14.7 | 8.6 |
| Hodgkin Lymphoma | 2.8 | 1.5 | 1.4 | 0.4 | [l] | 0.5 | [l] | 0.9 | 1.1 | 0.5 | 1.0 | 0.5 | 2.5 | 0.7 | 3.0 | 1.9 | 4.2 | 1.7 | 3.1 | 0.8 | 2.7 | 1.5 |
| Non-Hodgkin Lymphoma | 13.3 | 12.8 | 9.3 | 4.3 | 7.7 | 6.4 | 8.5 | 9.6 | 10.5 | 4.3 | 7.9 | 5.2 | 9.6 | 2.7 | 10.7 | 6.3 | 17.2 | 6.4 | 12.0 | 4.7 | 11.0 | 5.2 |
| Multiple myeloma | 3.6 | 3.8 | 1.6 | 0.9 | 0.6 | 2.5 | 1.6 | 1.6 | 3.1 | 0.8 | 2.2 | 1.0 | 4.1 | 1.1 | 2.9 | 2.1 | 7.4 | 1.7 | 7.5 | 1.1 | 2.6 | 1.9 |
| Leukemia | 10.3 | 8.0 | 6.8 | 5.1 | 2.1 | 5.8 | 5.8 | 6.1 | 6.5 | 5.7 | 5.9 | 5.5 | 7.8 | 3.7 | 10.7 | 6.5 | 9.1 | 5.7 | 7.5 | 2.8 | 9.3 | 5.4 |

Source: Adapted from Statistics Canada, Canadian Census Health and Environment Cohorts 2006 & 2011, 2006 long-form census, 2011 National Household Survey, Canadian Vital Statistics Death Database 2006-2015, and Canadian Cancer Registry 2006-2015; and from GLOBOCAN.[1]

CanCHECs=Canadian Census Health and Environment Cohorts; CNS=central nervous system.

[a] GLOBOCAN 2020 estimates for China.

[b] GLOBOCAN 2020 estimates for Republic of Korea.
[c] GLOBOCAN 2020 estimates for Japan.
[d] GLOBOCAN 2020 estimates for Philippines.
[e] GLOBOCAN 2020 estimates for South-Eastern Asia.
[f] GLOBOCAN 2020 estimates for South-Central Asia.
[g] The definition of "Western Asian" in the 2006 long-form census and 2011 National Household survey included some countries (Afghanistan, Iran) defined as South-Central Asia by GLOBOCAN 2020. The CanCHECs estimates should therefore be compared with both adjacent columns of GLOBOCAN 2020 estimates from South-Central Asia & Western Asia.
[h] GLOBOCAN 2020 estimates for Western Asia.
[i] GLOBOCAN 2020 estimates for Northern Africa and Western Asia (combined).
[j] GLOBOCAN 2020 estimates for Eastern Africa, Middle Africa, Southern Africa, and Western Africa (combined).
[k] GLOBOCAN 2020 estimates for Latin America and the Caribbean.
[l] Number of cases under disclosure threshold for cancer site.

### Supplementary Figures

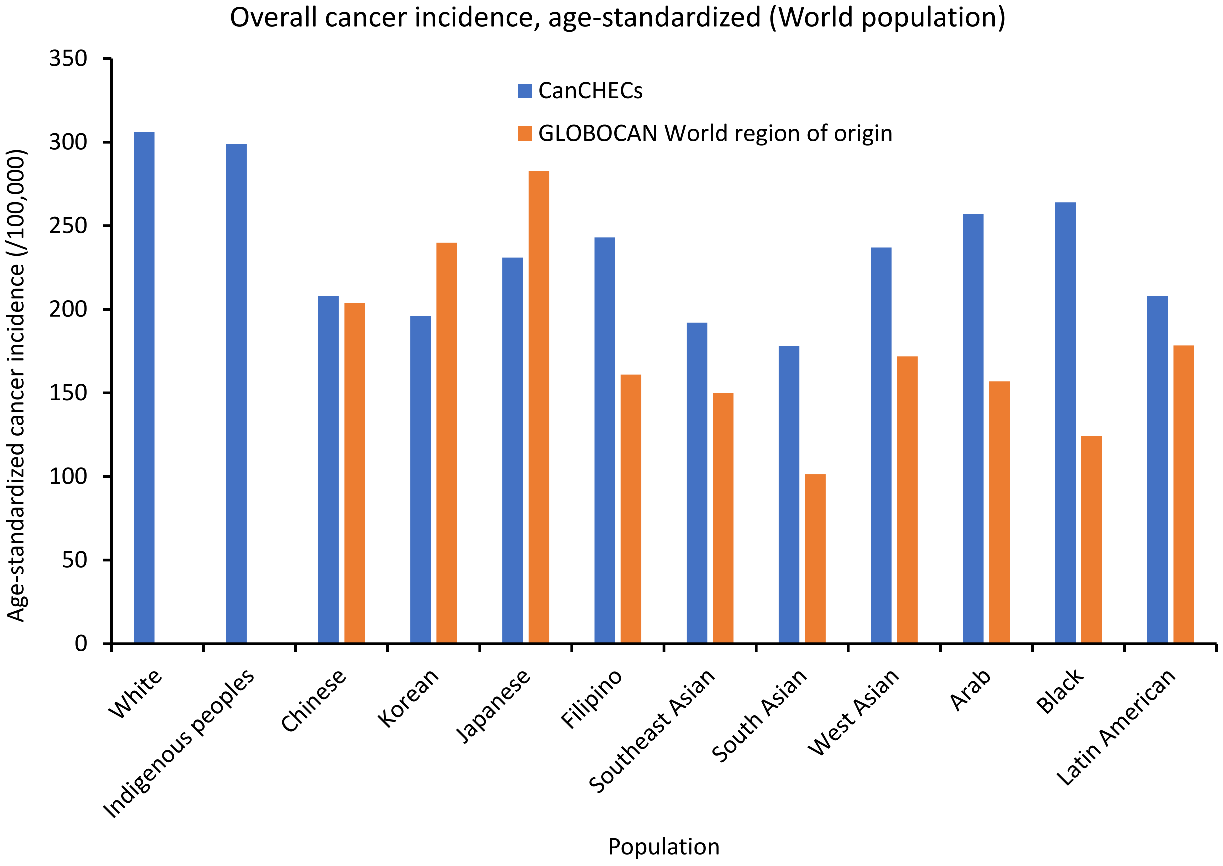

Supplementary Figure 1. Age-standardized incidence rate of cancer per 100,000 by racial group, all cancer sites combined. Standardized to the 1960 Segi World population. Source: Adapted from: Statistics Canada, Canadian Census Health and Environment Cohorts 2006 & 2011, 2006 long-form census, 2011 National Household Survey, Canadian Vital Statistics Death Database 2006-2015, and Canadian Cancer Registry 2006-2015; and from GLOBOCAN 2020.[1] Definitions for world region of origin used for each group can be found in the footnotes of Supplementary Table 4. CanCHECs=Canadian Census Health and Environment Cohorts.

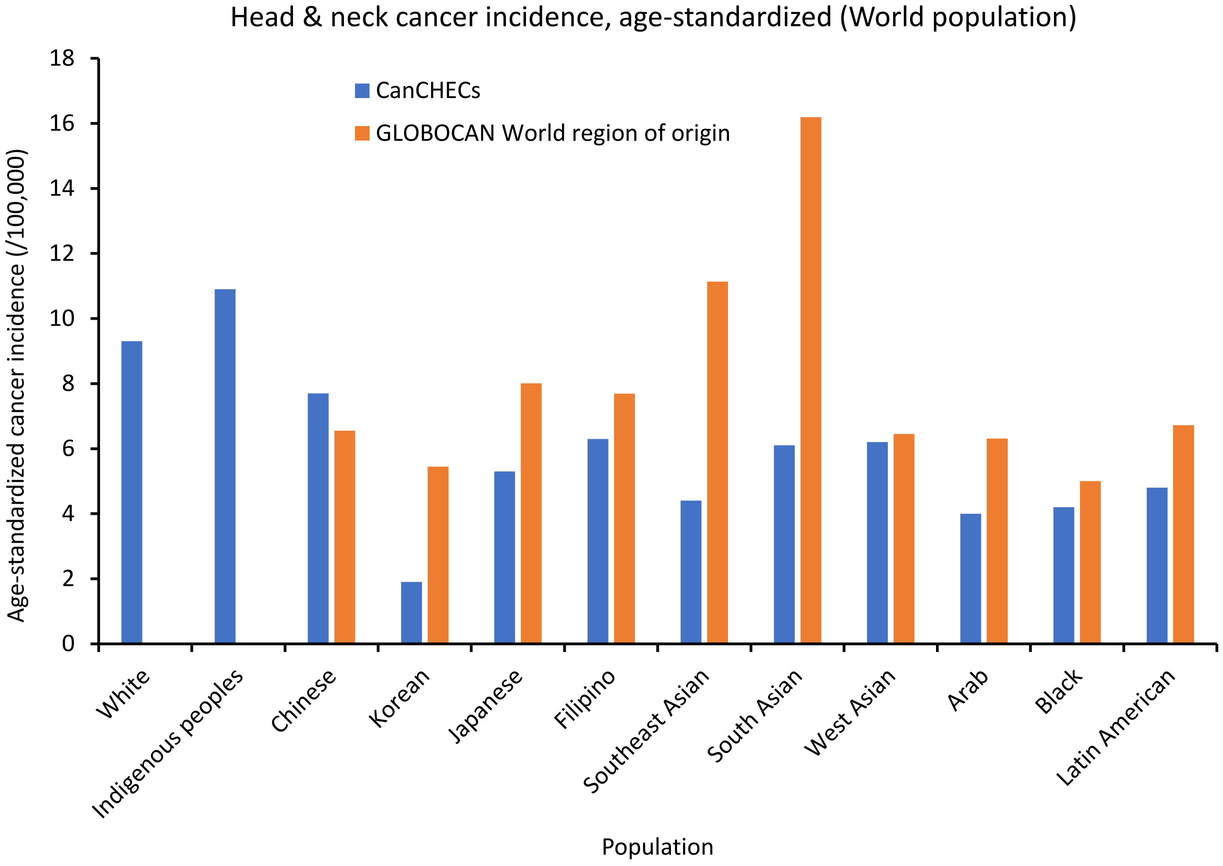

Supplementary Figure 2. Age-standardized incidence rate of head and neck cancers per 100,000 by racial group. Standardized to the 1960 Segi World population. Source: Adapted from: Statistics Canada, Canadian Census Health and Environment Cohorts 2006 & 2011, 2006 long-form census, 2011 National Household Survey, Canadian Vital Statistics Death Database 2006-2015, and Canadian Cancer Registry 2006-2015; and from GLOBOCAN 2020.[1] Definitions for world region of origin used for each group can be found in the footnotes of Supplementary Table 4. CanCHECs=Canadian Census Health and Environment Cohorts.

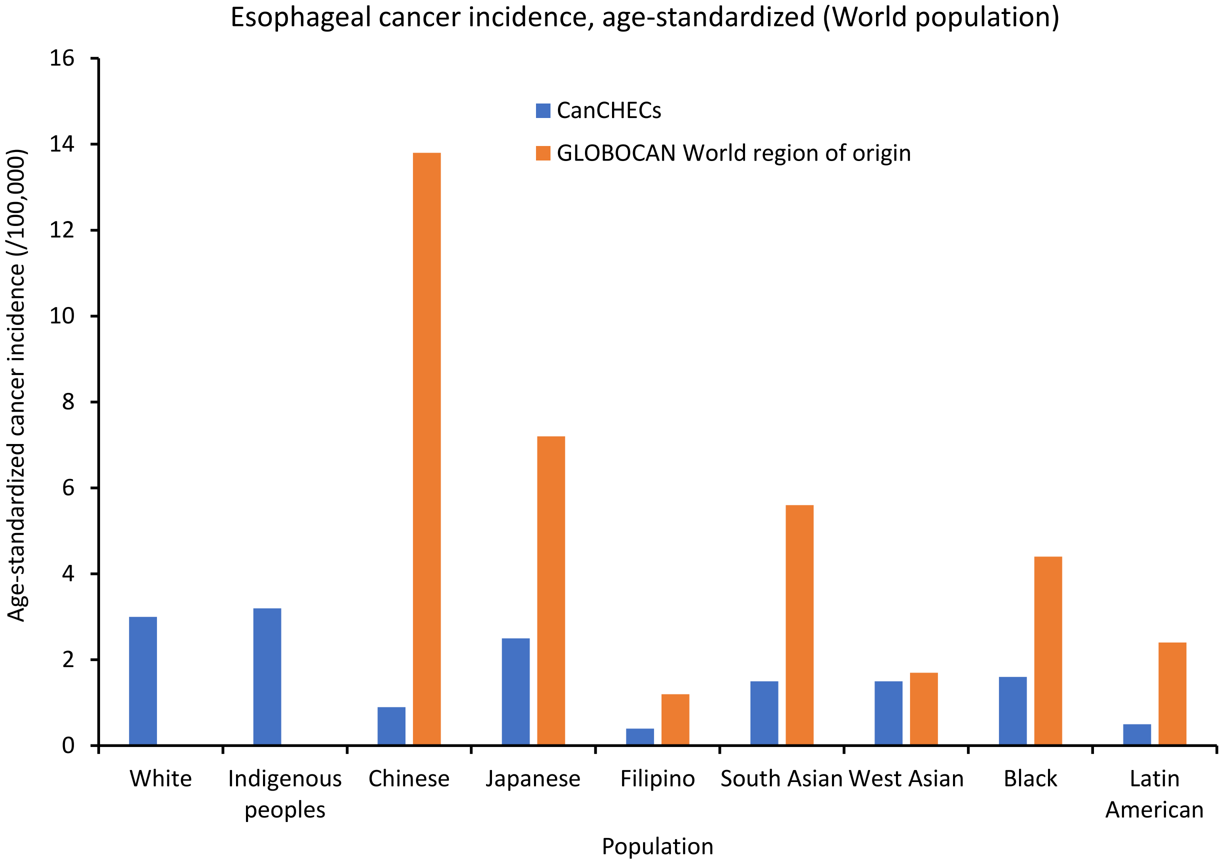

Supplementary Figure 3. Age-standardized incidence rate of esophageal cancers per 100,000 by racial group. Standardized to the 1960 Segi World population. Source: Adapted from: Statistics Canada, Canadian Census Health and Environment Cohorts 2006 & 2011, 2006 long-form census, 2011 National Household Survey, Canadian Vital Statistics Death Database 2006-2015, and Canadian Cancer Registry 2006-2015; and from GLOBOCAN 2020.[1] Definitions for world region of origin used for each group can be found in the footnotes of Supplementary Table 4. CanCHECs=Canadian Census Health and Environment Cohorts.

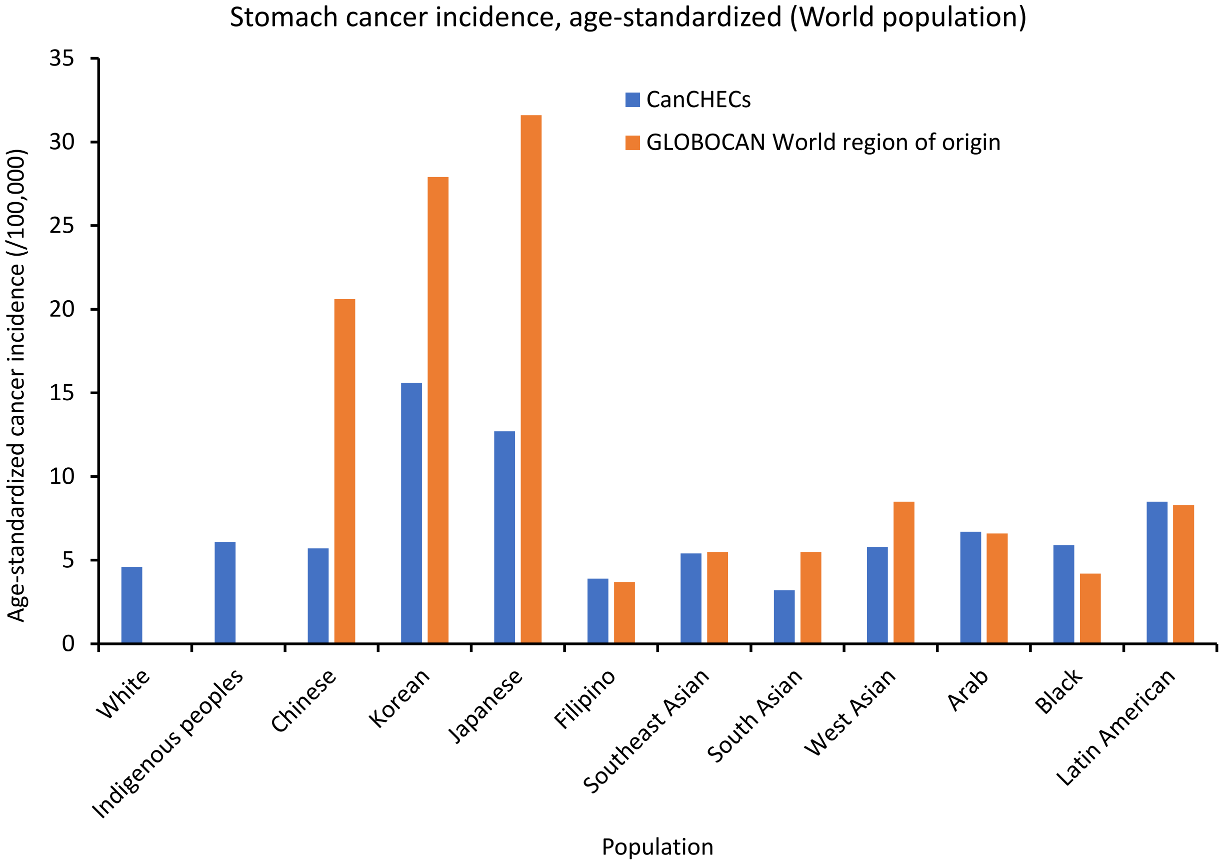

Supplementary Figure 4. Age-standardized incidence rate of stomach cancers per 100,000 by racial group. Standardized to the 1960 Segi World population. Source: Adapted from: Statistics Canada, Canadian Census Health and Environment Cohorts 2006 & 2011, 2006 long-form census, 2011 National Household Survey, Canadian Vital Statistics Death Database 2006-2015, and Canadian Cancer Registry 2006-2015; and from GLOBOCAN 2020.[1] Definitions for world region of origin used for each group can be found in the footnotes of Supplementary Table 4. CanCHECs=Canadian Census Health and Environment Cohorts.

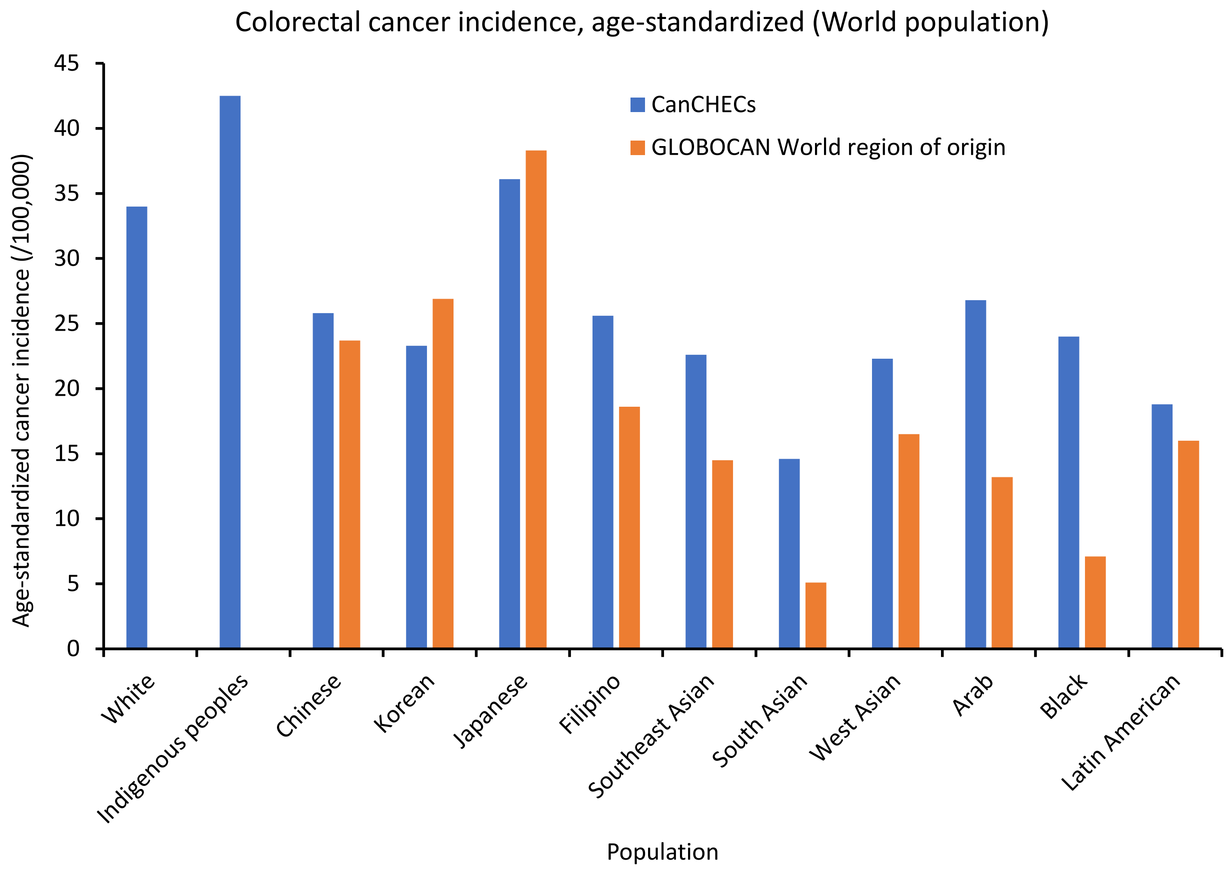

Supplementary Figure 5. Age-standardized incidence rate of colorectal cancers per 100,000 by racial group. Standardized to the 1960 Segi World population. Source: Adapted from: Statistics Canada, Canadian Census Health and Environment Cohorts 2006 & 2011, 2006 long-form census, 2011 National Household Survey, Canadian Vital Statistics Death Database 2006-2015, and Canadian Cancer Registry 2006-2015; and from GLOBOCAN 2020.[1] Definitions for world region of origin used for each group can be found in the footnotes of Supplementary Table 4. CanCHECs=Canadian Census Health and Environment Cohorts.

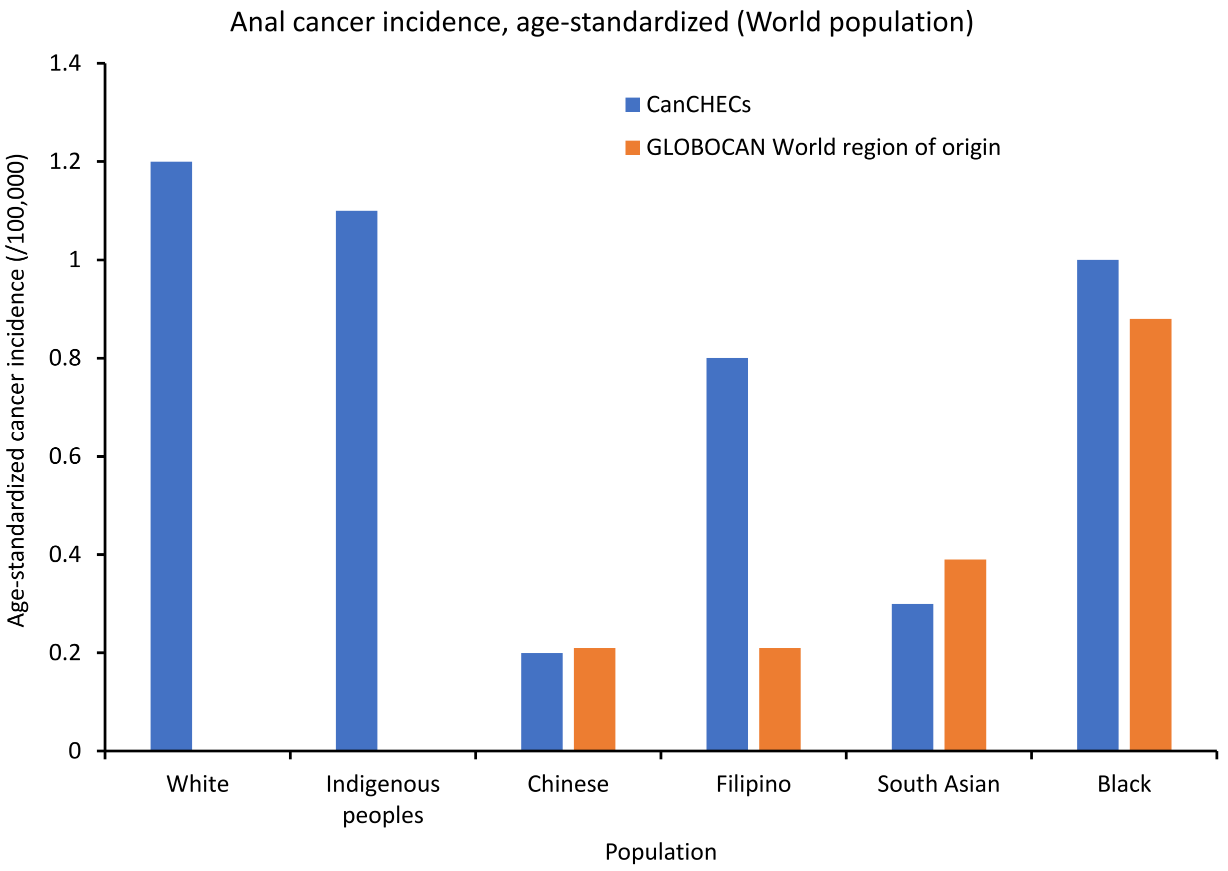

Supplementary Figure 6. Age-standardized incidence rate of anal cancers per 100,000 by racial group. Standardized to the 1960 Segi World population. Source: Adapted from: Statistics Canada, Canadian Census Health and Environment Cohorts 2006 & 2011, 2006 long-form census, 2011 National Household Survey, Canadian Vital Statistics Death Database 2006-2015, and Canadian Cancer Registry 2006-2015; and from GLOBOCAN 2020.[1] Definitions for world region of origin used for each group can be found in the footnotes of Supplementary Table 4. CanCHECs=Canadian Census Health and Environment Cohorts.

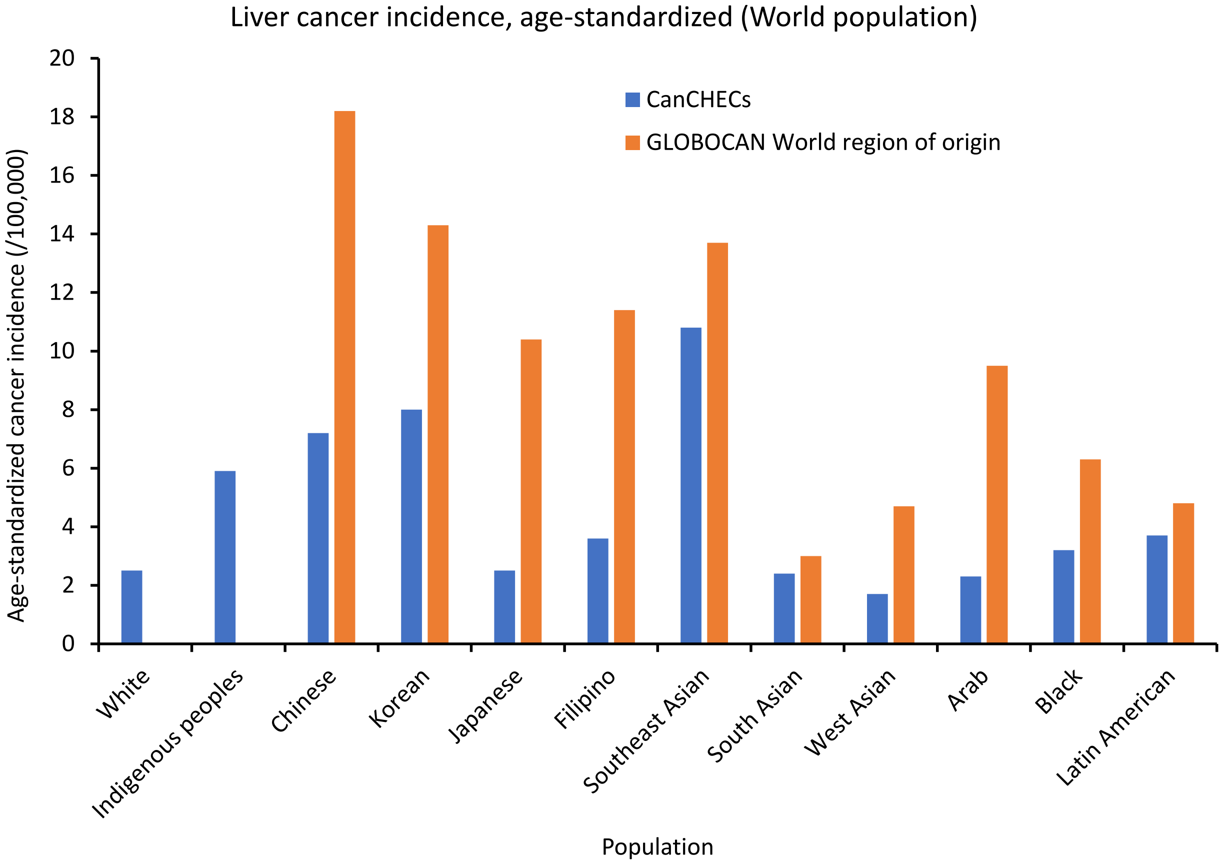

Supplementary Figure 7. Age-standardized incidence rate of liver cancers per 100,000 by racial group. Standardized to the 1960 Segi World population. Source: Adapted from: Statistics Canada, Canadian Census Health and Environment Cohorts 2006 & 2011, 2006 long-form census, 2011 National Household Survey, Canadian Vital Statistics Death Database 2006-2015, and Canadian Cancer Registry 2006-2015; and from GLOBOCAN 2020.[1] Definitions for world region of origin used for each group can be found in the footnotes of Supplementary Table 4. CanCHECs=Canadian Census Health and Environment Cohorts.

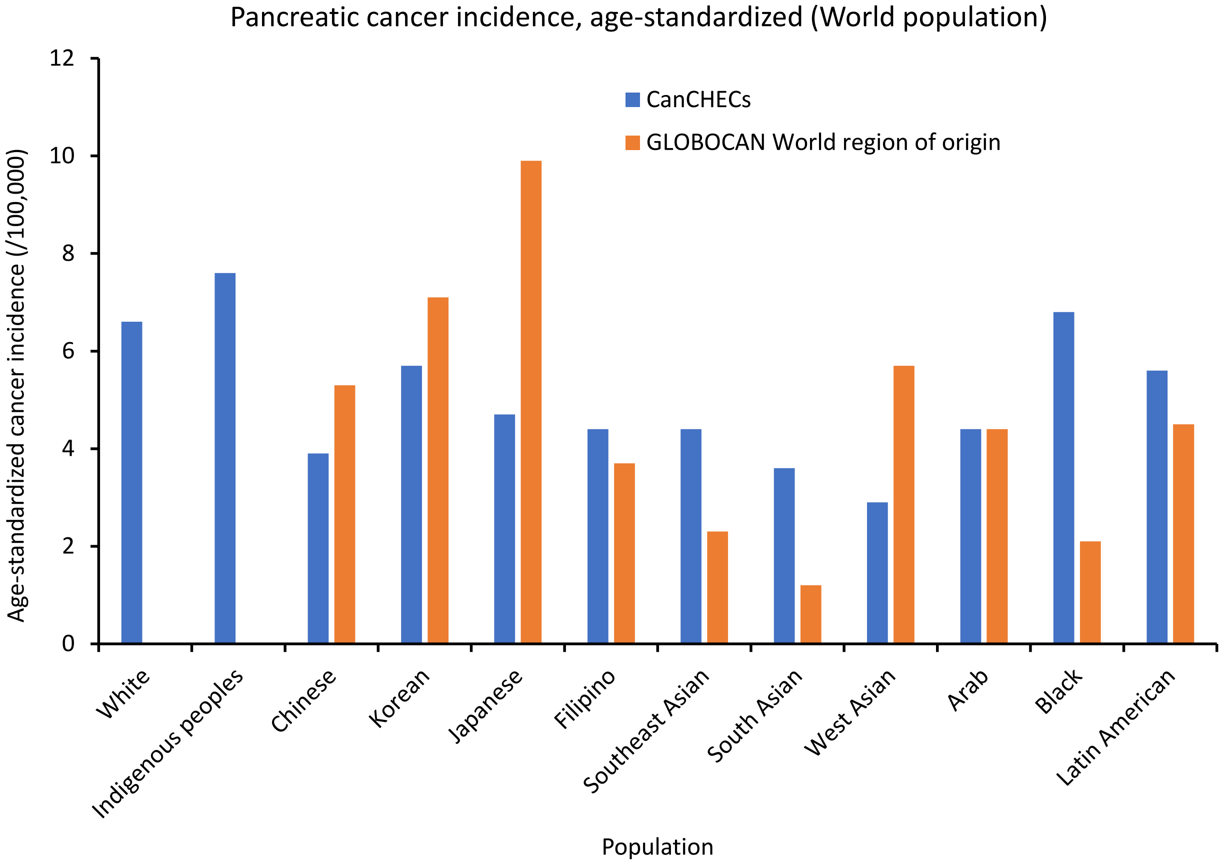

Supplementary Figure 8. Age-standardized incidence rate of pancreatic cancers per 100,000 by racial group. Standardized to the 1960 Segi World population. Source: Adapted from: Statistics Canada, Canadian Census Health and Environment Cohorts 2006 & 2011, 2006 long-form census, 2011 National Household Survey, Canadian Vital Statistics Death Database 2006-2015, and Canadian Cancer Registry 2006-2015; and from GLOBOCAN 2020.[1] Definitions for world region of origin used for each group can be found in the footnotes of Supplementary Table 4. CanCHECs=Canadian Census Health and Environment Cohorts.

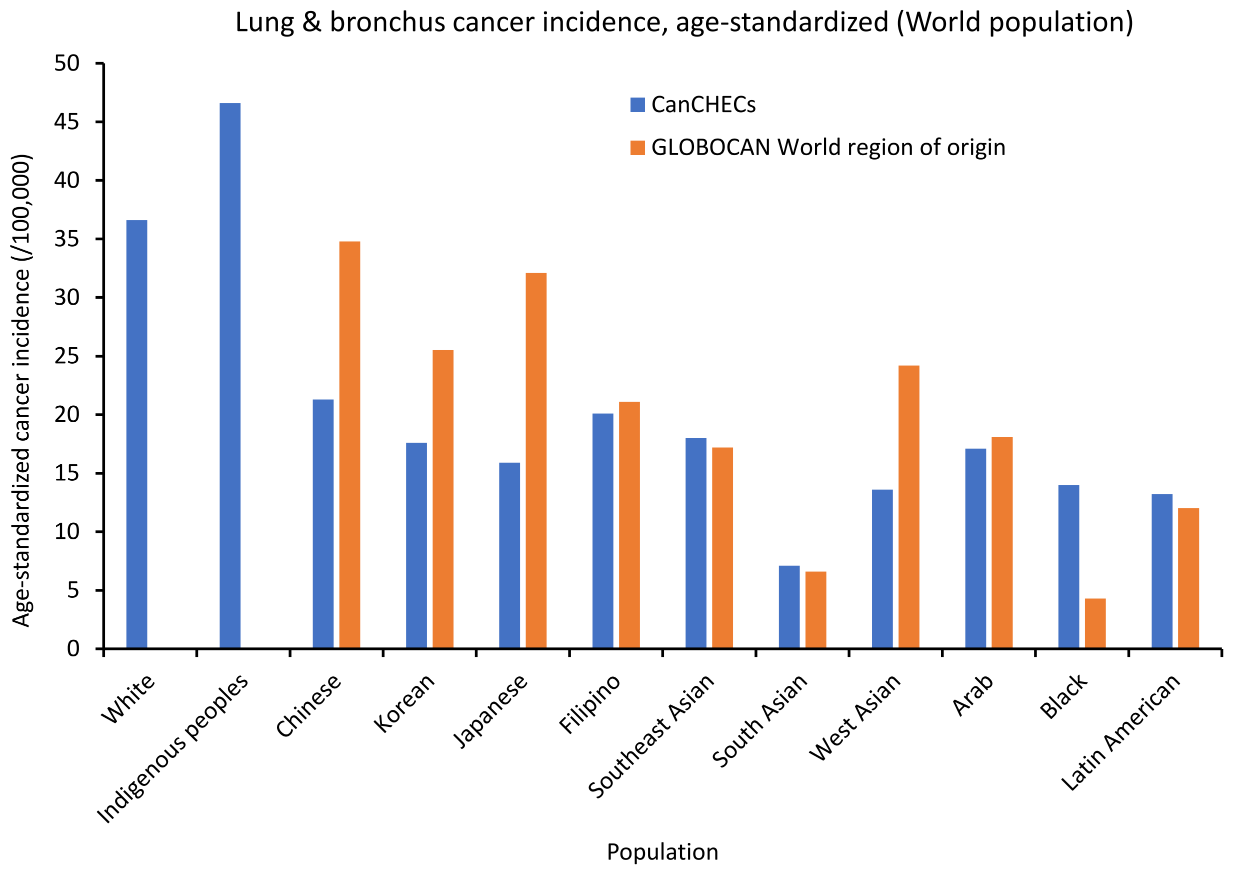

Supplementary Figure 9. Age-standardized incidence rate of lung and bronchus cancers per 100,000 by racial group. Standardized to the 1960 Segi World population. Source: Adapted from: Statistics Canada, Canadian Census Health and Environment Cohorts 2006 & 2011, 2006 long-form census, 2011 National Household Survey, Canadian Vital Statistics Death Database 2006-2015, and Canadian Cancer Registry 2006-2015; and from GLOBOCAN 2020.[1] Definitions for world region of origin used for each group can be found in the footnotes of Supplementary Table 4. CanCHECs=Canadian Census Health and Environment Cohorts.

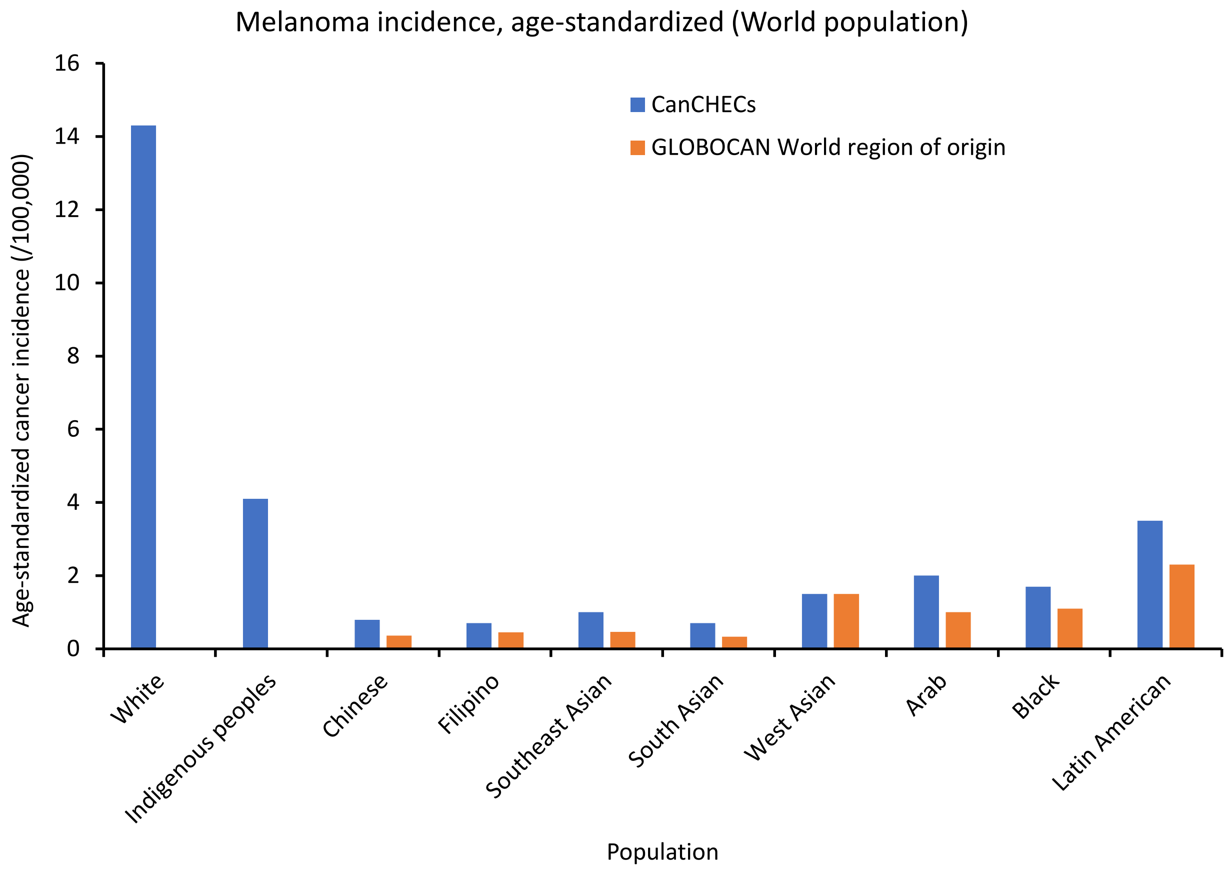

Supplementary Figure 10. Age-standardized incidence rate of melanomas per 100,000 by racial group. Standardized to the 1960 Segi World population. Source: Adapted from: Statistics Canada, Canadian Census Health and Environment Cohorts 2006 & 2011, 2006 long-form census, 2011 National Household Survey, Canadian Vital Statistics Death Database 2006-2015, and Canadian Cancer Registry 2006-2015; and from GLOBOCAN 2020.[1] Definitions for world region of origin used for each group can be found in the footnotes of Supplementary Table 4. CanCHECs=Canadian Census Health and Environment Cohorts.

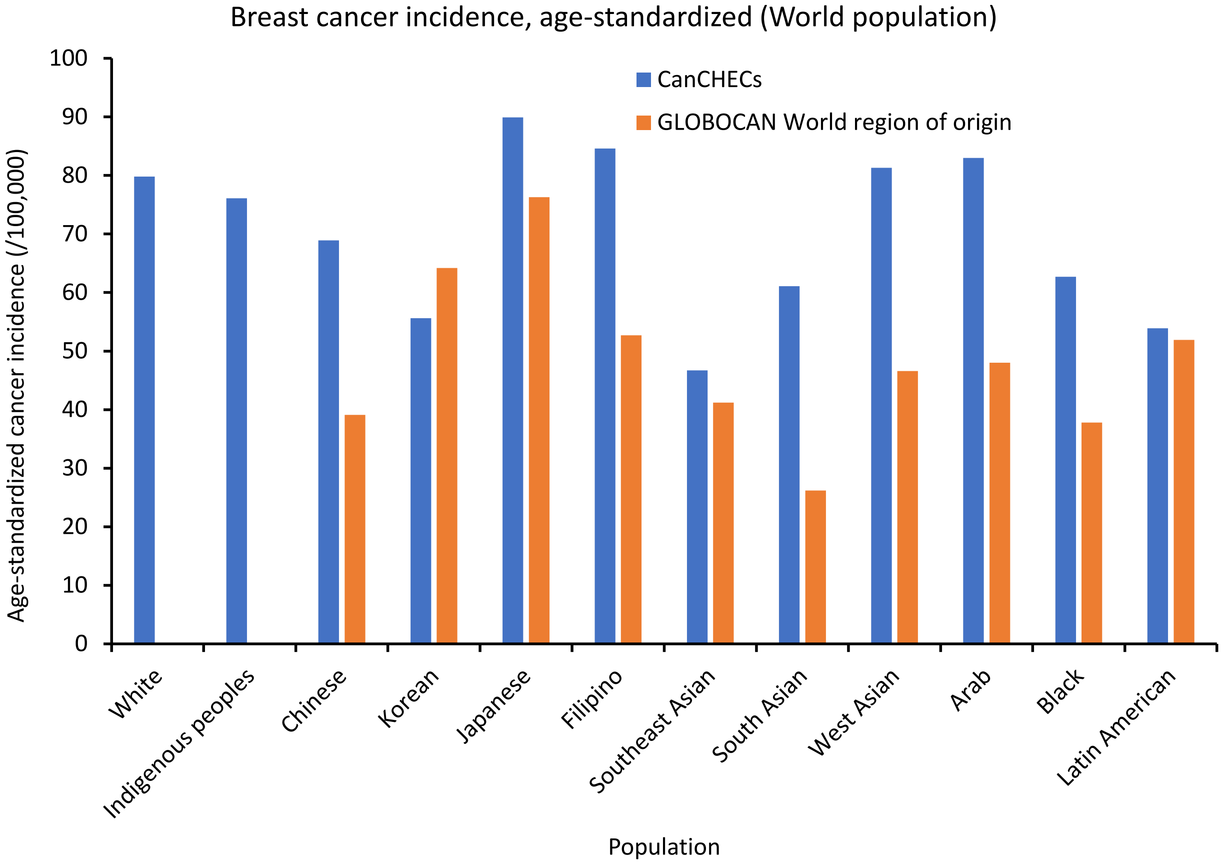

Supplementary Figure 11. Age-standardized incidence rate of breast cancer in females per 100,000 by racial group. Standardized to the 1960 Segi World population. Source: Adapted from: Statistics Canada, Canadian Census Health and Environment Cohorts 2006 & 2011, 2006 long-form census, 2011 National Household Survey, Canadian Vital Statistics Death Database 2006-2015, and Canadian Cancer Registry 2006-2015; and from GLOBOCAN 2020.[1] Definitions for world region of origin used for each group can be found in the footnotes of Supplementary Table 4. CanCHECs=Canadian Census Health and Environment Cohorts.

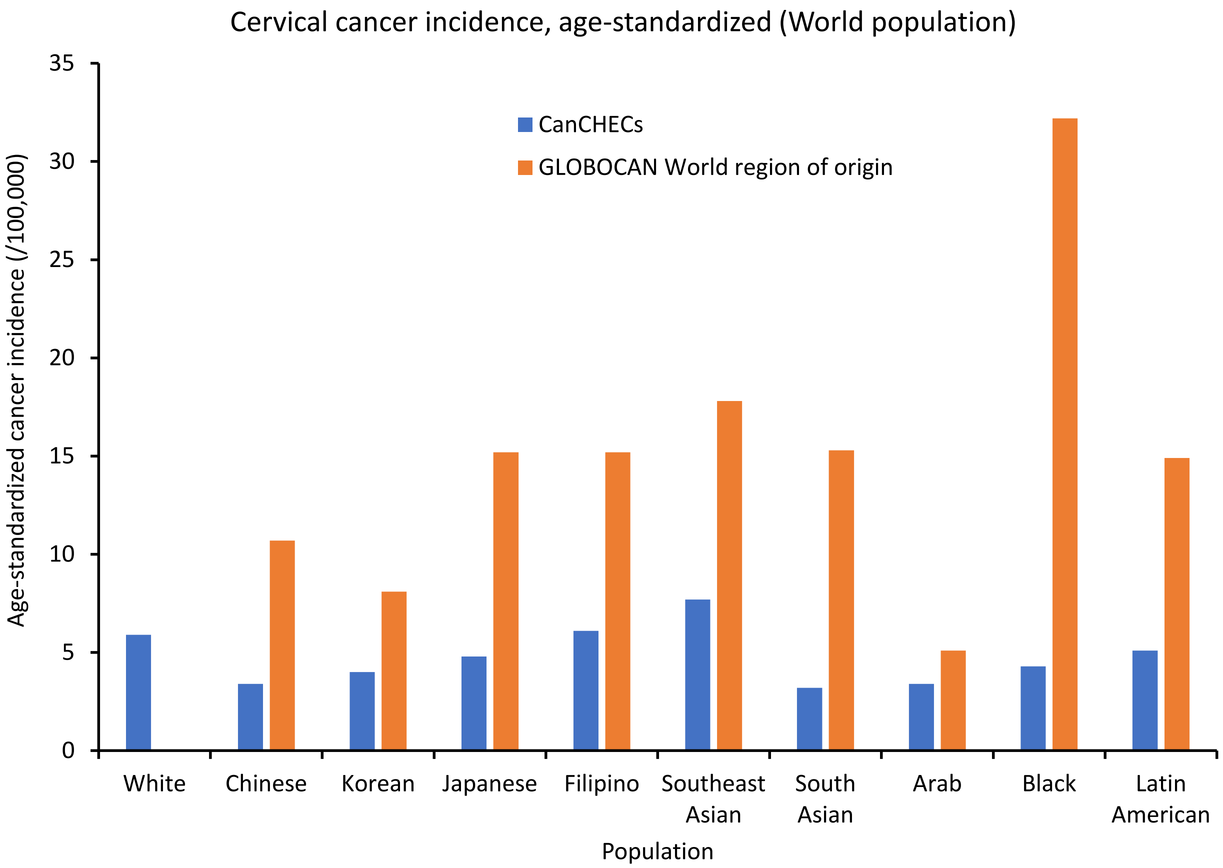

Supplementary Figure 12. Age-standardized incidence rate of cervical cancer in females per 100,000 by racial group. Standardized to the 1960 Segi World population. Source: Adapted from: Statistics Canada, Canadian Census Health and Environment Cohorts 2006 & 2011, 2006 long-form census, 2011 National Household Survey, Canadian Vital Statistics Death Database 2006-2015, and Canadian Cancer Registry 2006-2015; and from GLOBOCAN 2020.[1] Definitions for world region of origin used for each group can be found in the footnotes of Supplementary Table 4. CanCHECs=Canadian Census Health and Environment Cohorts.

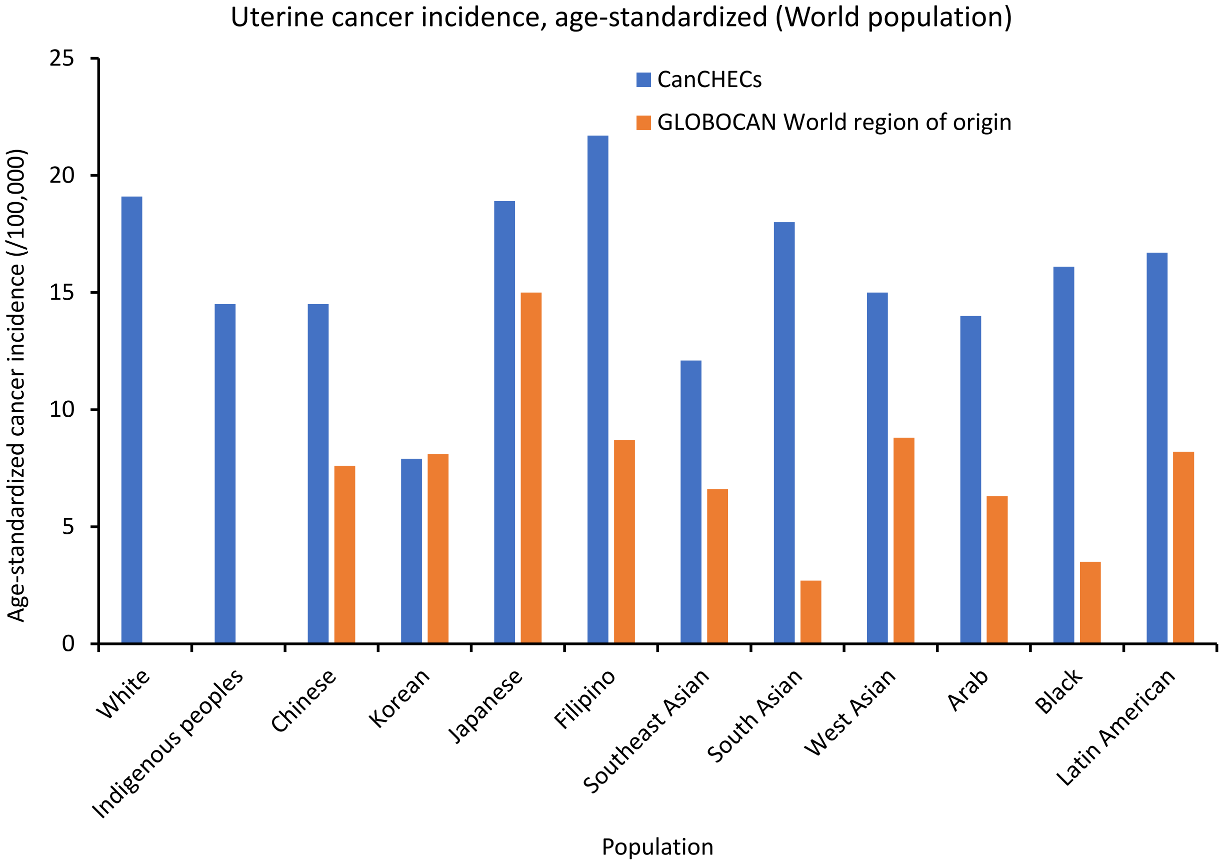

Supplementary Figure 13. Age-standardized incidence rate of uterine cancer in females per 100,000 by racial group. Standardized to the 1960 Segi World population. Source: Adapted from: Statistics Canada, Canadian Census Health and Environment Cohorts 2006 & 2011, 2006 long-form census, 2011 National Household Survey, Canadian Vital Statistics Death Database 2006-2015, and Canadian Cancer Registry 2006-2015; and from GLOBOCAN 2020.[1] Definitions for world region of origin used for each group can be found in the footnotes of Supplementary Table 4. CanCHECs=Canadian Census Health and Environment Cohorts.

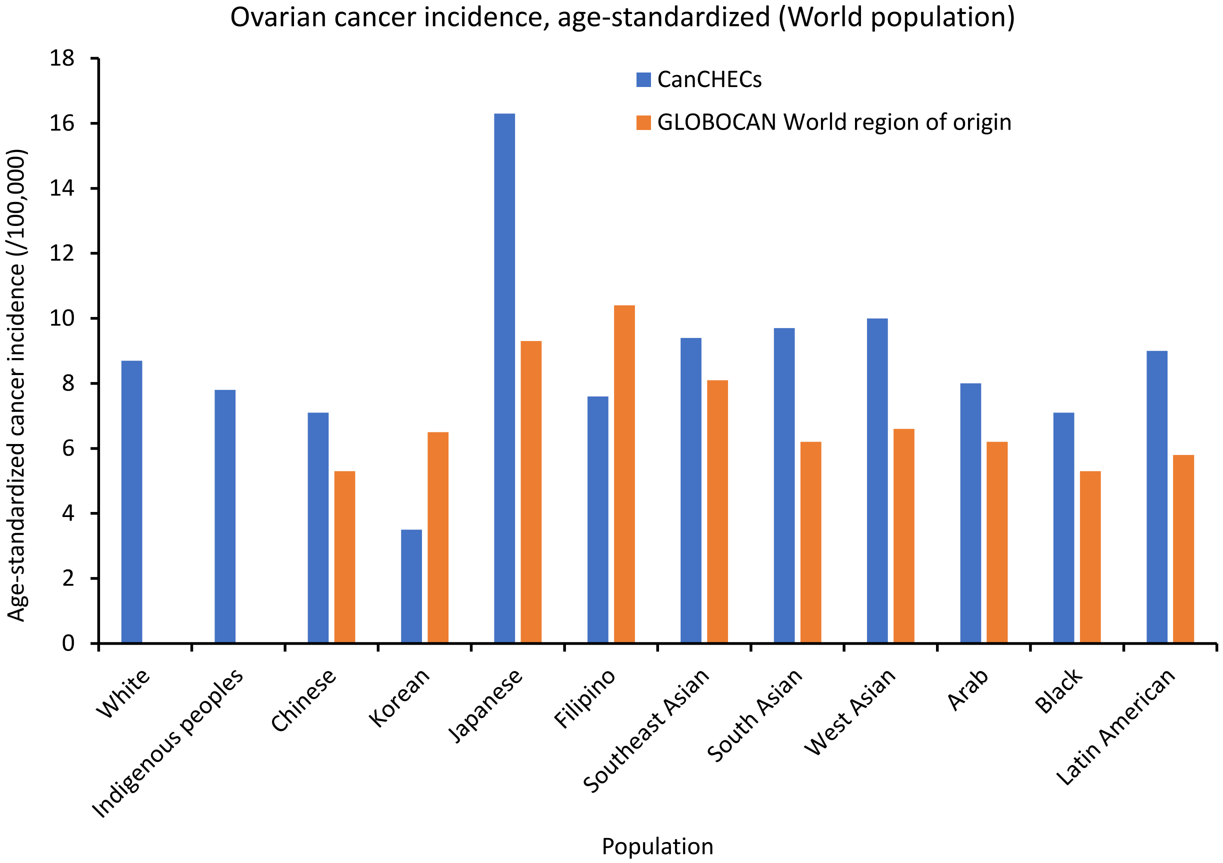

Supplementary Figure 14. Age-standardized incidence rate of ovarian cancer in females per 100,000 by racial group. Standardized to the 1960 Segi World population. Source: Adapted from: Statistics Canada, Canadian Census Health and Environment Cohorts 2006 & 2011, 2006 long-form census, 2011 National Household Survey, Canadian Vital Statistics Death Database 2006-2015, and Canadian Cancer Registry 2006-2015; and from GLOBOCAN 2020.[1] Definitions for world region of origin used for each group can be found in the footnotes of Supplementary Table 4. CanCHECs=Canadian Census Health and Environment Cohorts.

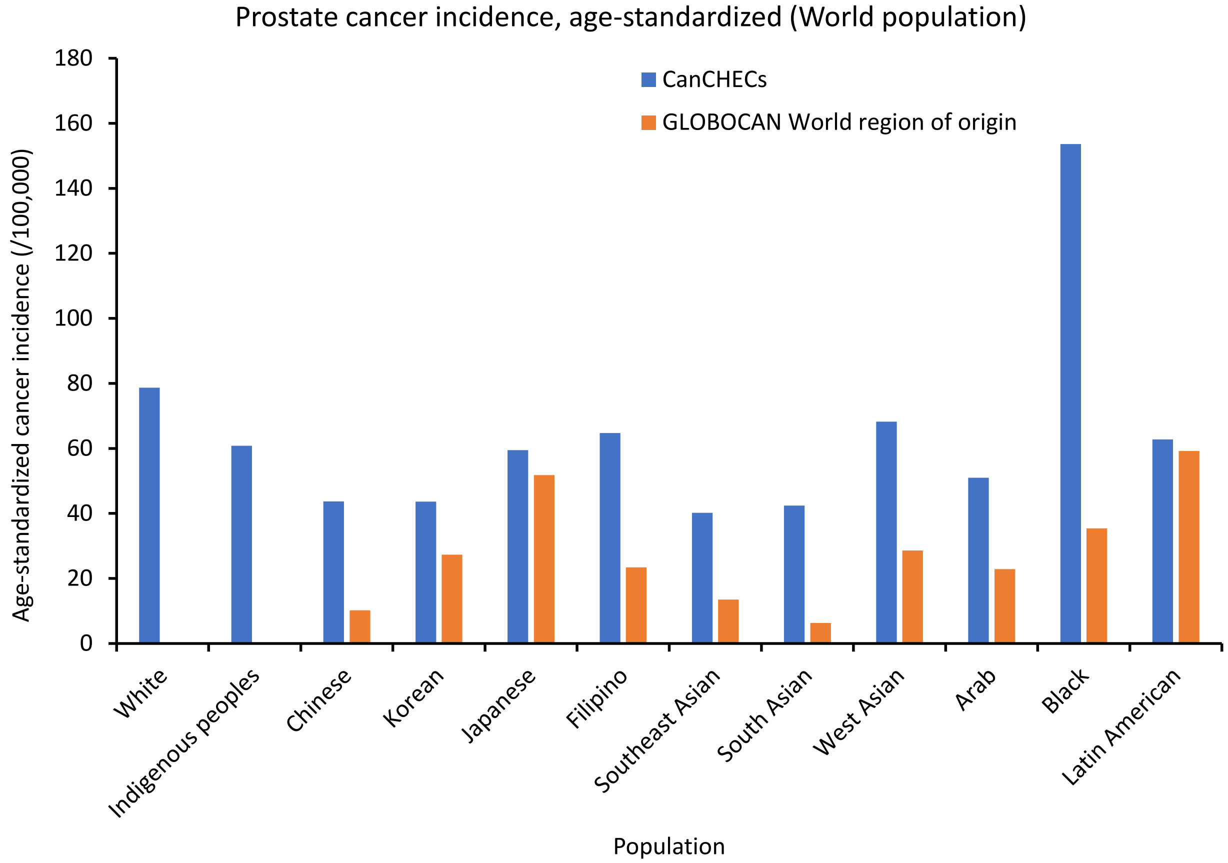

Supplementary Figure 15. Age-standardized incidence rate of prostate cancer in males per 100,000 by racial group. Standardized to the 1960 Segi World population. Source: Adapted from: Statistics Canada, Canadian Census Health and Environment Cohorts 2006 & 2011, 2006 long-form census, 2011 National Household Survey, Canadian Vital Statistics Death Database 2006-2015, and Canadian Cancer Registry 2006-2015; and from GLOBOCAN 2020.[1] Definitions for world region of origin used for each group can be found in the footnotes of Supplementary Table 4. CanCHECs=Canadian Census Health and Environment Cohorts.

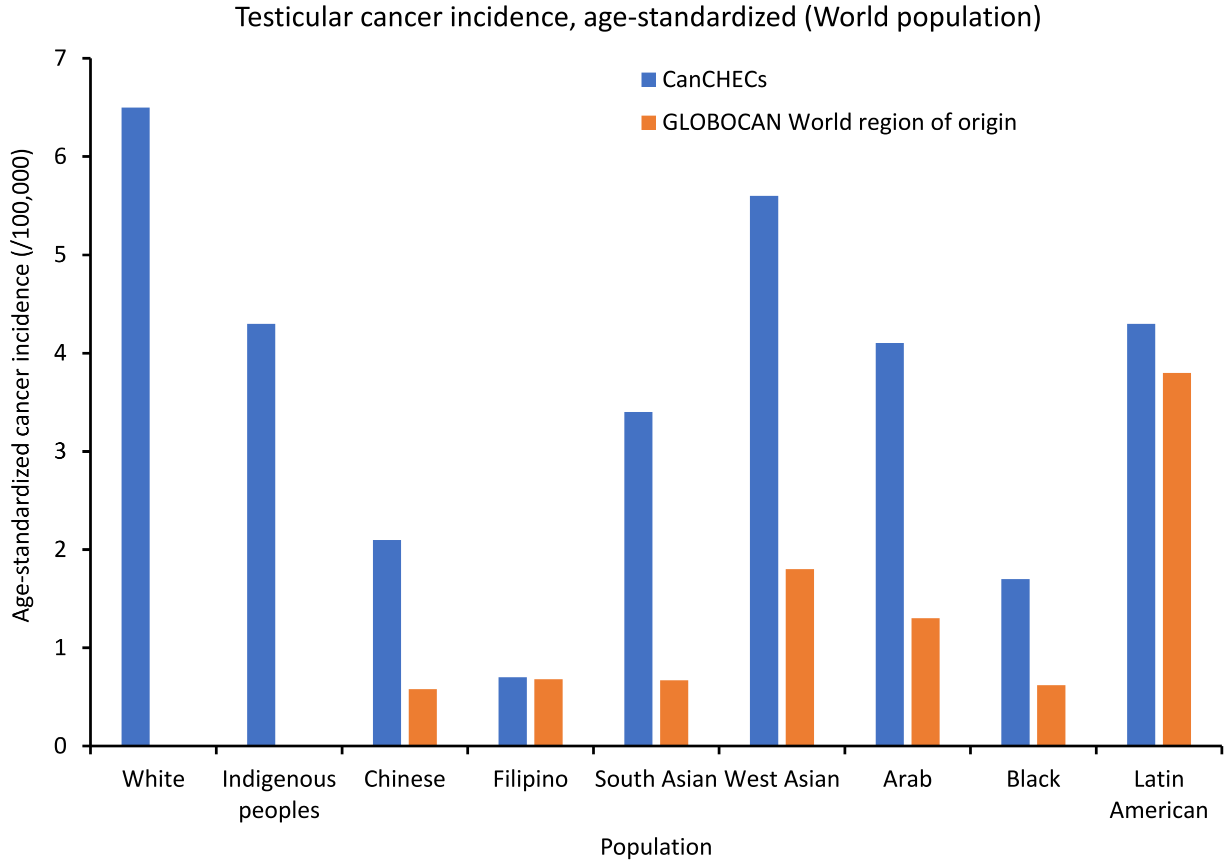

Supplementary Figure 16. Age-standardized incidence rate of testicular cancer in males per 100,000 by racial group. Standardized to the 1960 Segi World population. Source: Adapted from: Statistics Canada, Canadian Census Health and Environment Cohorts 2006 & 2011, 2006 long-form census, 2011 National Household Survey, Canadian Vital Statistics Death Database 2006-2015, and Canadian Cancer Registry 2006-2015; and from GLOBOCAN 2020.[1] Definitions for world region of origin used for each group can be found in the footnotes of Supplementary Table 4. CanCHECs=Canadian Census Health and Environment Cohorts.

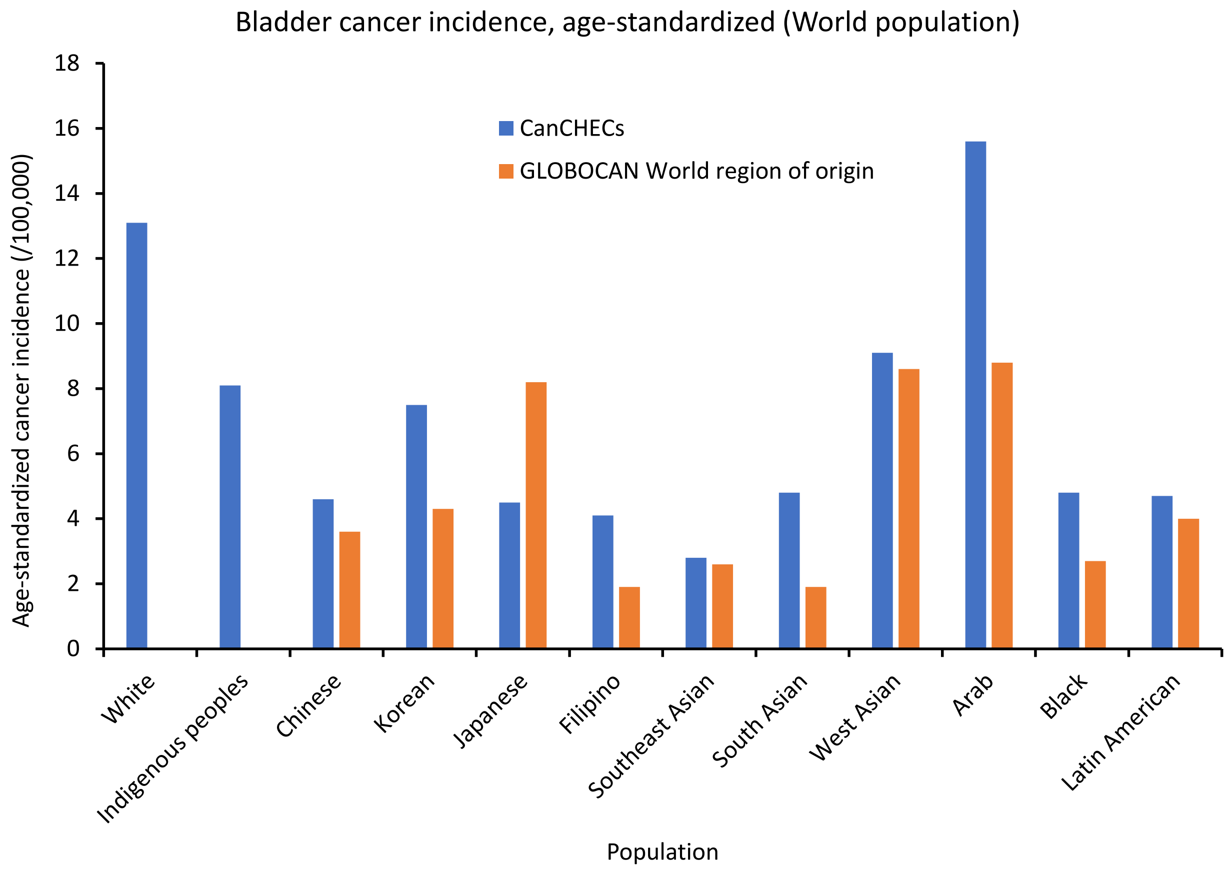

Supplementary Figure 17. Age-standardized incidence rate of bladder cancer per 100,000 by racial group. Standardized to the 1960 Segi World population. Source: Adapted from: Statistics Canada, Canadian Census Health and Environment Cohorts 2006 & 2011, 2006 long-form census, 2011 National Household Survey, Canadian Vital Statistics Death Database 2006-2015, and Canadian Cancer Registry 2006-2015; and from GLOBOCAN 2020.[1] Definitions for world region of origin used for each group can be found in the footnotes of Supplementary Table 4. CanCHECs=Canadian Census Health and Environment Cohorts.

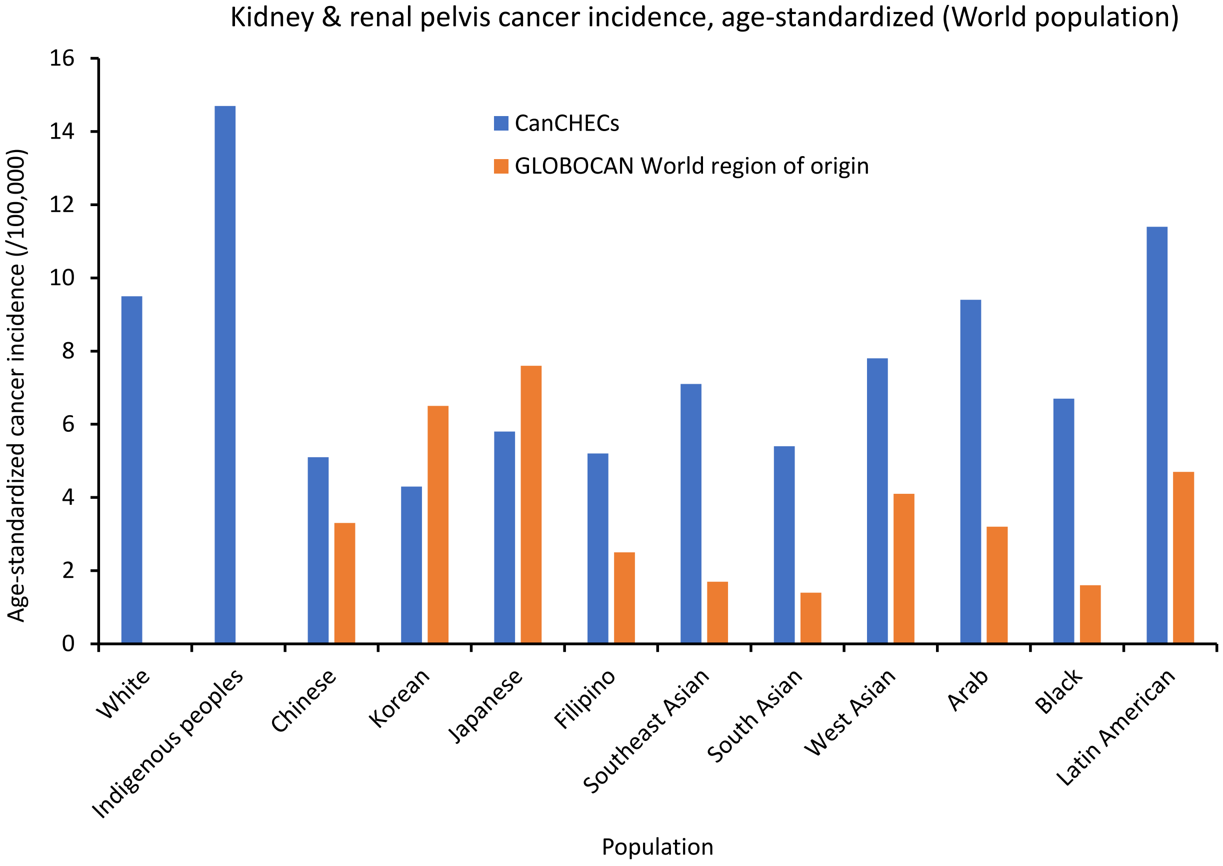

Supplementary Figure 18. Age-standardized incidence rate of kidney and renal pelvis cancer per 100,000 by racial group. Standardized to the 1960 Segi World population. Source: Adapted from: Statistics Canada, Canadian Census Health and Environment Cohorts 2006 & 2011, 2006 long-form census, 2011 National Household Survey, Canadian Vital Statistics Death Database 2006-2015, and Canadian Cancer Registry 2006-2015; and from GLOBOCAN 2020.[1] Definitions for world region of origin used for each group can be found in the footnotes of Supplementary Table 4. CanCHECs=Canadian Census Health and Environment Cohorts.

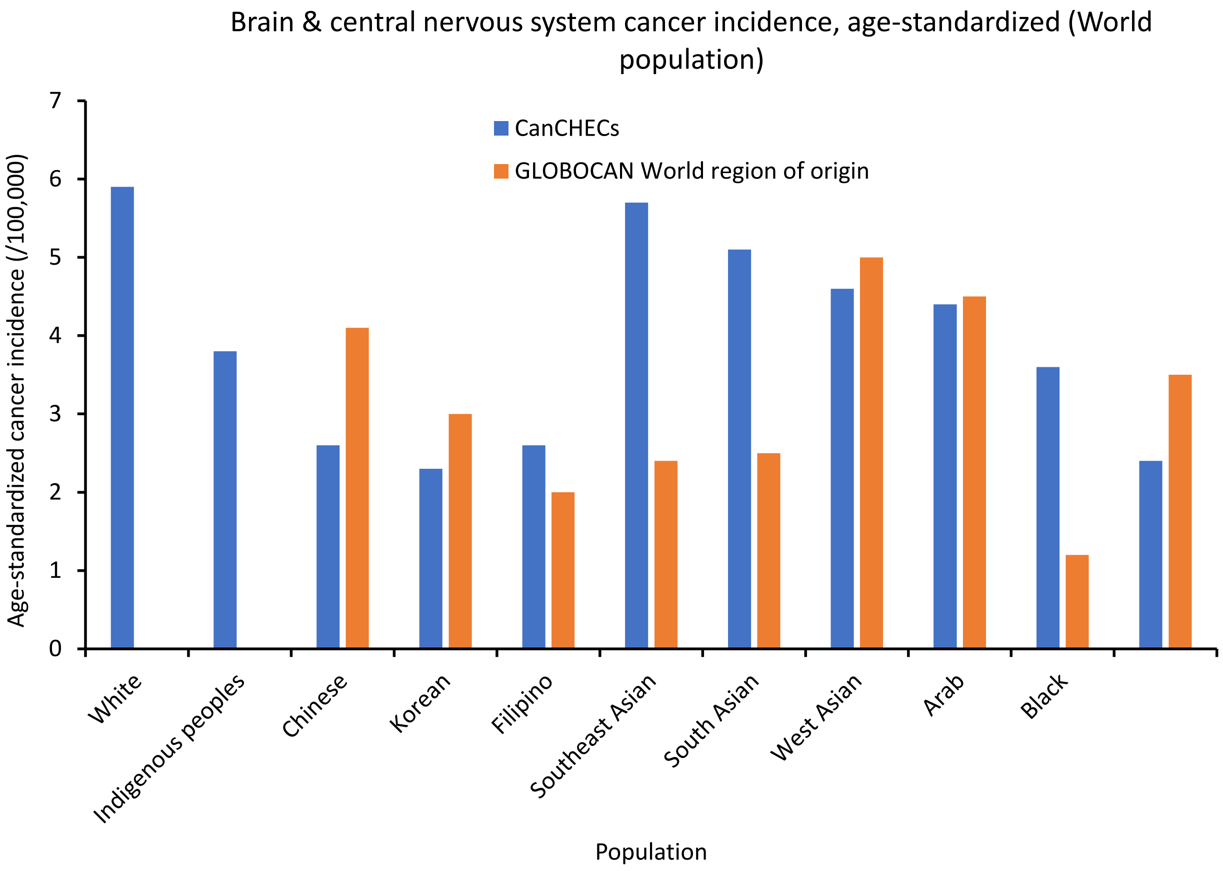

Supplementary Figure 19. Age-standardized incidence rate of brain & central nervous system cancers per 100,000 by racial group. Standardized to the 1960 Segi World population. Source: Adapted from: Statistics Canada, Canadian Census Health and Environment Cohorts 2006 & 2011, 2006 long-form census, 2011 National Household Survey, Canadian Vital Statistics Death Database 2006-2015, and Canadian Cancer Registry 2006-2015; and from GLOBOCAN 2020.[1] Definitions for world region of origin used for each group can be found in the footnotes of Supplementary Table 4. CanCHECs=Canadian Census Health and Environment Cohorts.

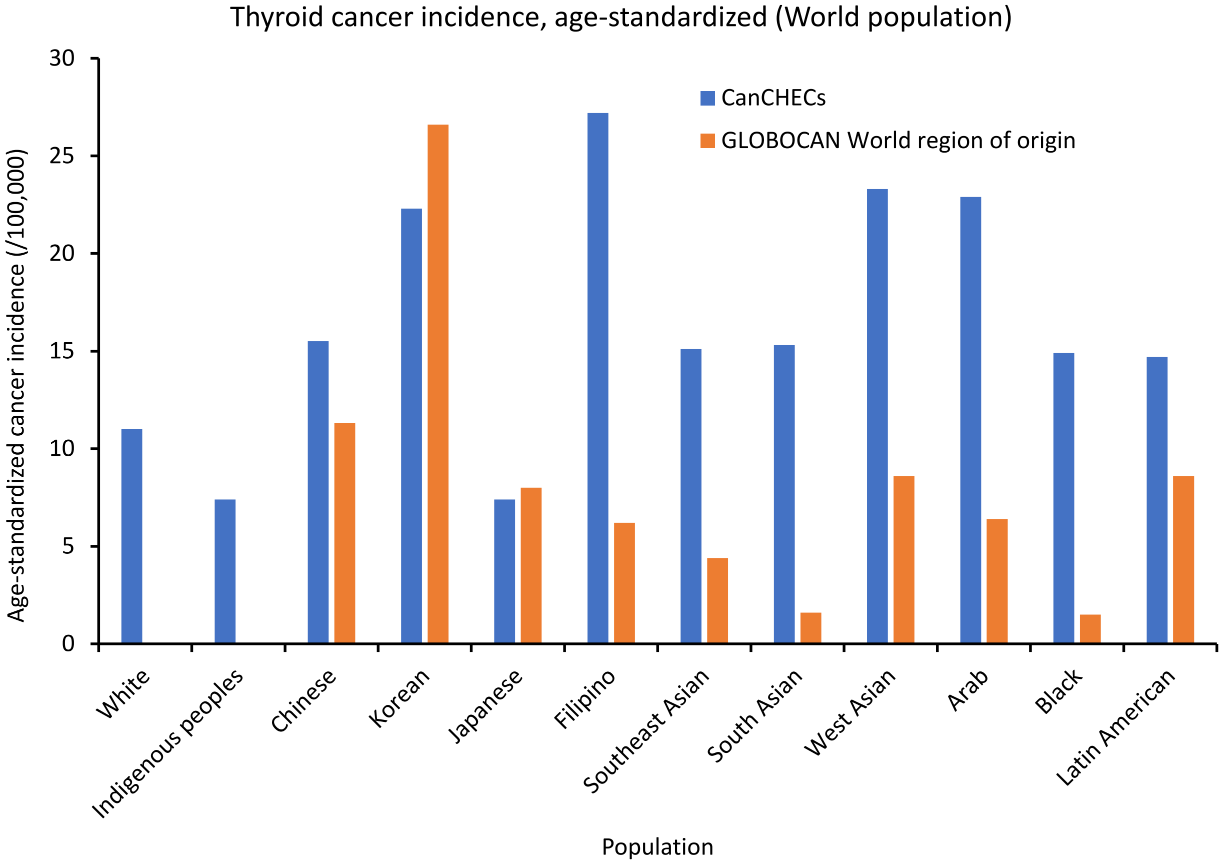

Supplementary Figure 20. Age-standardized incidence rate of thyroid cancers per 100,000 by racial group. Standardized to the 1960 Segi World population. Source: Adapted from: Statistics Canada, Canadian Census Health and Environment Cohorts 2006 & 2011, 2006 long-form census, 2011 National Household Survey, Canadian Vital Statistics Death Database 2006-2015, and Canadian Cancer Registry 2006-2015; and from GLOBOCAN 2020.[1] Definitions for world region of origin used for each group can be found in the footnotes of Supplementary Table 4. CanCHECs=Canadian Census Health and Environment Cohorts.

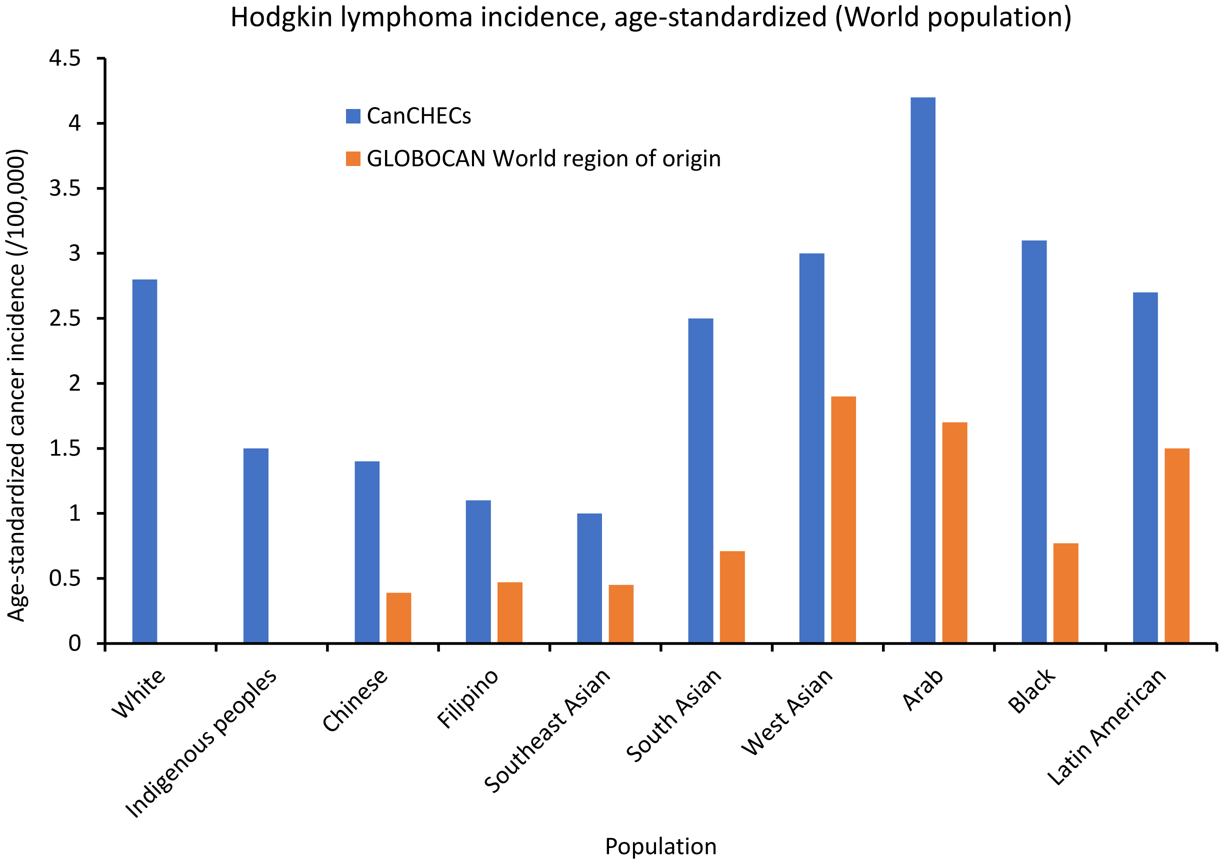

Supplementary Figure 21. Age-standardized incidence rate of Hodgkin lymphomas per 100,000 by racial group. Standardized to the 1960 Segi World population. Source: Adapted from: Statistics Canada, Canadian Census Health and Environment Cohorts 2006 & 2011, 2006 long-form census, 2011 National Household Survey, Canadian Vital Statistics Death Database 2006-2015, and Canadian Cancer Registry 2006-2015; and from GLOBOCAN 2020.[1] Definitions for world region of origin used for each group can be found in the footnotes of Supplementary Table 4. CanCHECs=Canadian Census Health and Environment Cohorts.

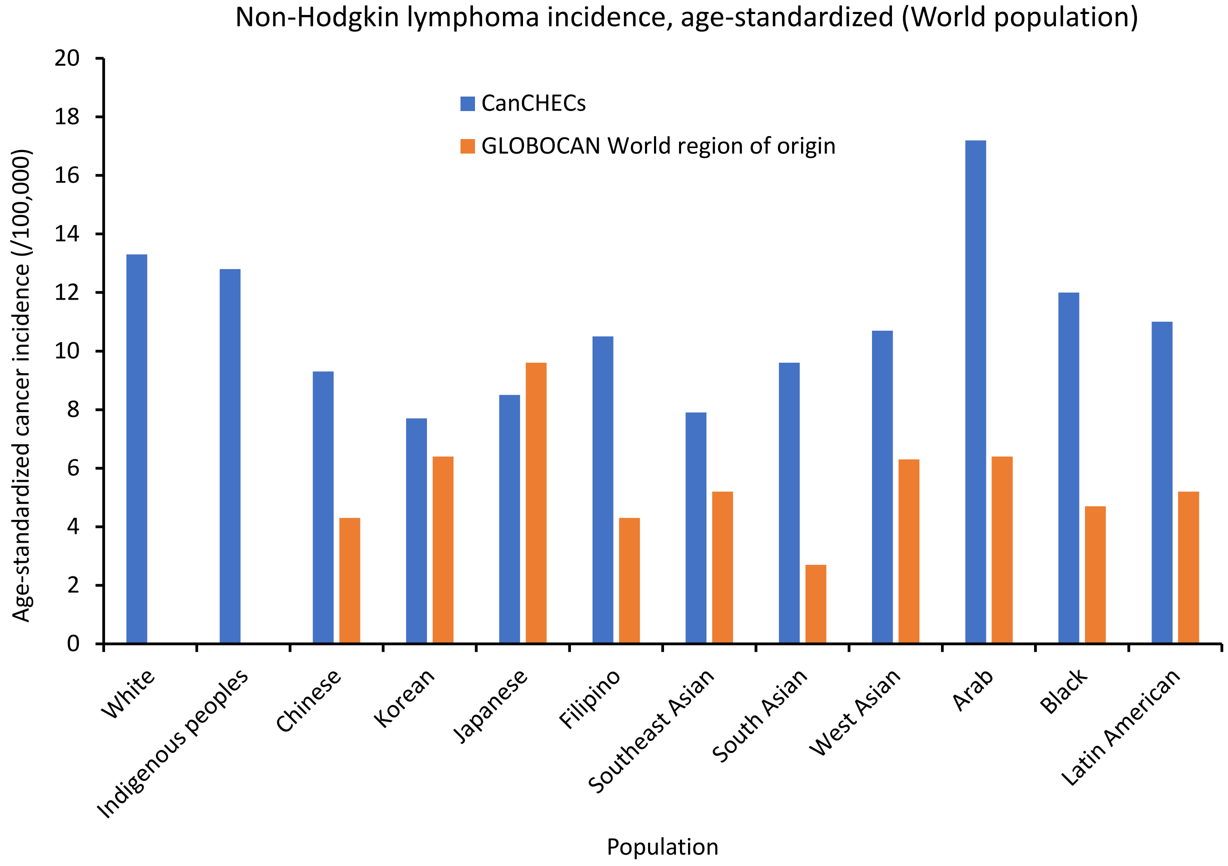

Supplementary Figure 22. Age-standardized incidence rate of non-Hodgkin lymphomas per 100,000 by racial group. Standardized to the 1960 Segi World population. Source: Adapted from: Statistics Canada, Canadian Census Health and Environment Cohorts 2006 & 2011, 2006 long-form census, 2011 National Household Survey, Canadian Vital Statistics Death Database 2006-2015, and Canadian Cancer Registry 2006-2015; and from GLOBOCAN 2020.[1] Definitions for world region of origin used for each group can be found in the footnotes of Supplementary Table 4. CanCHECs=Canadian Census Health and Environment Cohorts.

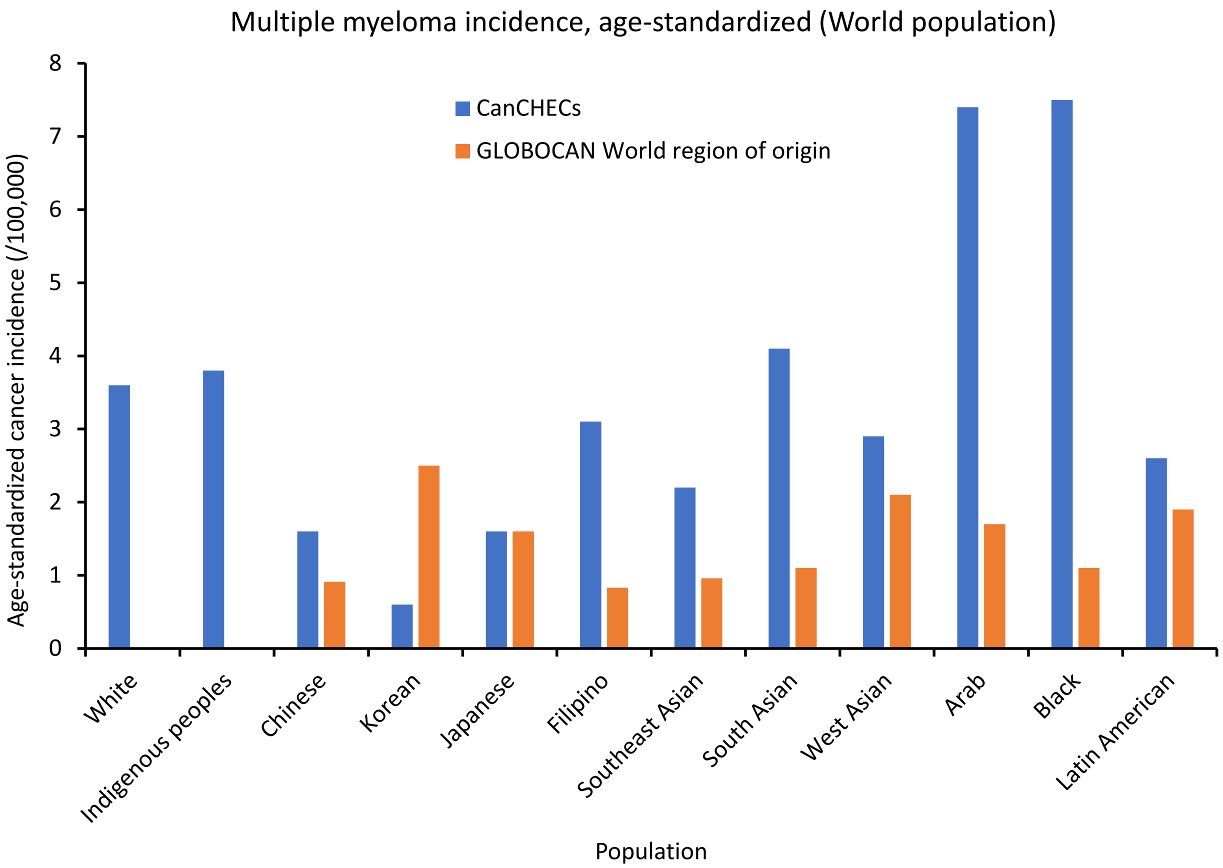

Supplementary Figure 23. Age-standardized incidence rate of multiple myelomas per 100,000 by racial group. Standardized to the 1960 Segi World population. Source: Adapted from: Statistics Canada, Canadian Census Health and Environment Cohorts 2006 & 2011, 2006 long-form census, 2011 National Household Survey, Canadian Vital Statistics Death Database 2006-2015, and Canadian Cancer Registry 2006-2015; and from GLOBOCAN 2020.[1] Definitions for world region of origin used for each group can be found in the footnotes of Supplementary Table 4. CanCHECs=Canadian Census Health and Environment Cohorts.

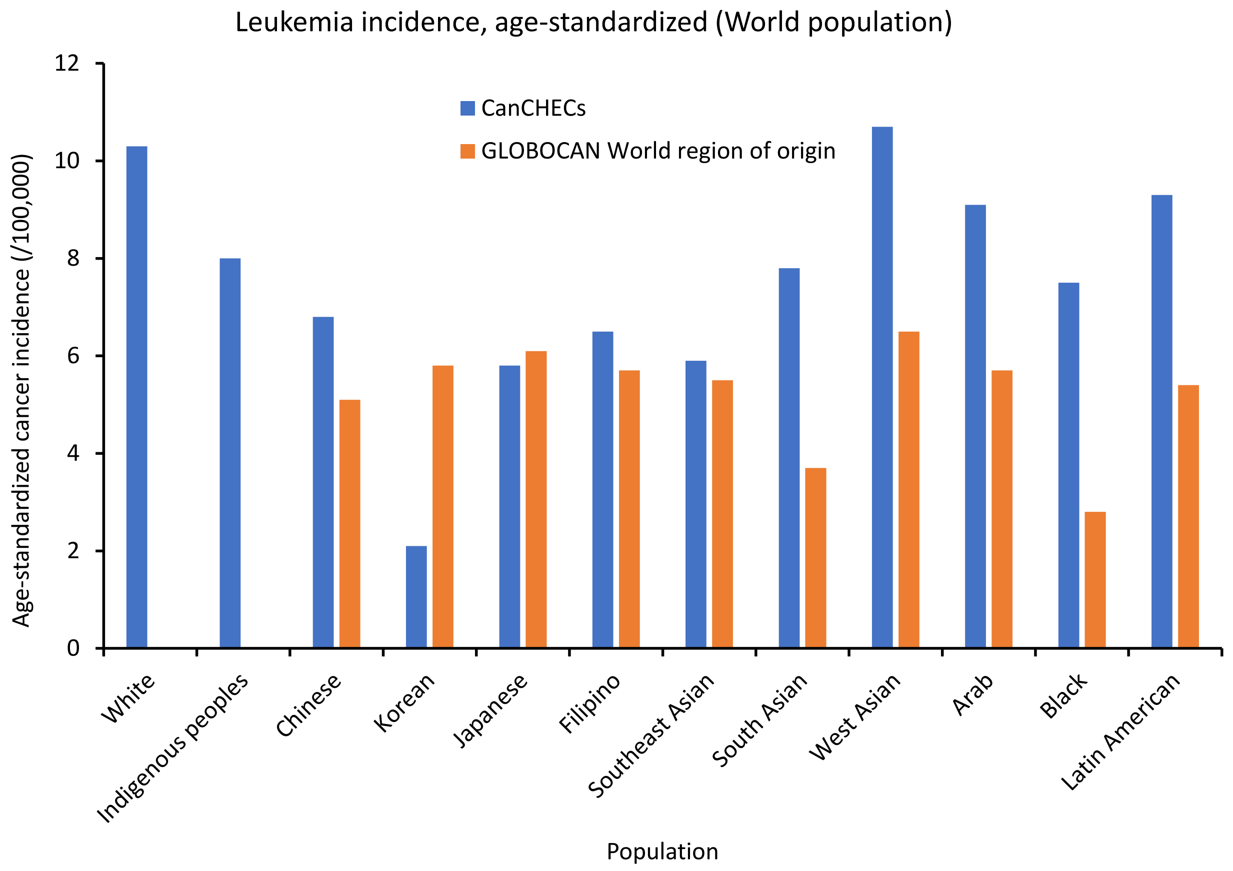

Supplementary Figure 24. Age-standardized incidence rate of leukemias per 100,000 by racial group. Standardized to the 1960 Segi World population. Source: Adapted from: Statistics Canada, Canadian Census Health and Environment Cohorts 2006 & 2011, 2006 long-form census, 2011 National Household Survey, Canadian Vital Statistics Death Database 2006-2015, and Canadian Cancer Registry 2006-2015; and from GLOBOCAN 2020.[1] Definitions for world region of origin used for each group can be found in the footnotes of Supplementary Table 4. CanCHECs=Canadian Census Health and Environment Cohorts.
